# Deleterious coding variation associated with autism is consistent across populations, as exemplified by admixed Latin American populations

**DOI:** 10.1101/2024.12.27.24319460

**Authors:** Marina Natividad Avila, Seulgi Jung, F. Kyle Satterstrom, Jack M. Fu, Tess Levy, Laura G. Sloofman, Lambertus Klei, Thariana Pichardo, Christine R. Stevens, Caroline M. Cusick, Jennifer L. Ames, Gabriele S. Campos, Hilda Cerros, Roberto Chaskel, Claudia I. S. Costa, Michael L. Cuccaro, Andrea del Pilar Lopez, Magdalena Fernandez, Eugenio Ferro, Liliana Galeano, Ana Cristina D. E. S. Girardi, Anthony J. Griswold, Luis C. Hernandez, Naila Lourenço, Yunin Ludena, Diana L. Nuñez, Rosa Oyama, Katherine P. Peña, Isaac Pessah, Rebecca Schmidt, Holly M. Sweeney, Lizbeth Tolentino, Jaqueline Y. T. Wang, Lilia Albores-Gallo, Lisa A. Croen, Carlos S. Cruz-Fuentes, Irva Hertz-Picciotto, Alexander Kolevzon, Maria C. Lattig, Liliana Mayo, Maria Rita Passos-Bueno, Margaret A. Pericak-Vance, Paige M. Siper, Flora Tassone, M. Pilar Trelles, Autism Sequencing Consortium, Michael E. Talkowski, Mark J. Daly, Behrang Mahjani, Silvia De Rubeis, Edwin H. Cook, Kathryn Roeder, Catalina Betancur, Bernie Devlin, Joseph D. Buxbaum

## Abstract

The past decade has seen remarkable progress in identifying genes that, when impacted by deleterious coding variation, confer high risk for autism spectrum disorder (ASD), intellectual disability, and other developmental disorders. However, most underlying gene discovery efforts have focused on individuals of European ancestry, limiting insights into genetic risks across diverse populations. To help address this, the Genomics of Autism in Latin American Ancestries Consortium (GALA) was formed, presenting here the largest sequencing study of ASD in Latin American individuals (n>15,000). We identified 35 genome-wide significant (FDR < 0.05) ASD risk genes, with substantial overlap with findings from European cohorts, and highly constrained genes showing consistent signal across populations. The results provide support for emerging (e.g., *MARK2*, *YWHAG*, *PACS1*, *RERE, SPEN, GSE1, GLS, TNPO3, ANKRD17*) and established ASD genes, and for the utility of genetic testing approaches for deleterious variants in diverse populations, while also demonstrating the ongoing need for more inclusive genetic research and testing. We conclude that the biology of ASD is universal and not impacted to any detectable degree by ancestry.

**Autism Sequencing Consortium (ASC):** Branko Aleksic, Mykyta Artomov, Mafalda Barbosa, Elisa Benetti, Catalina Betancur, Monica Biscaldi-Schafer, Anders D. Børglum, Harrison Brand, Alfredo Brusco, Joseph D. Buxbaum, Gabriele Campos, Simona Cardaropoli, Diana Carli, Angel Carracedo, Marcus C. Y. Chan, Andreas G. Chiocchetti, Brian H. Y. Chung, Brett Collins, Ryan L. Collins, Edwin H. Cook, Hilary Coon, Claudia I. S. Costa, Michael L. Cuccaro, David J. Cutler, Mark J. Daly, Silvia De Rubeis, Bernie Devlin, Ryan N. Doan, Enrico Domenici, Shan Dong, Chiara Fallerini, Magdalena Fernandez, Montserrat Fernández-Prieto, Giovanni Battista Ferrero, Eugenio Ferro, Jennifer Foss Feig, Christine M. Freitag, Jack M. Fu, Liliana Galeano, J. Jay Gargus, Sherif Gerges, Elisa Giorgio, Ana Cristina Girardi, Stephen Guter, Emily Hansen-Kiss, Erina Hara, Danielle Halpern, Gail E. Herman, Luis C. Hernandez, Irva Hertz-Picciotto, David M. Hougaard, Christina M. Hultman, Suma Jacob, Miia Kaartinen, Lambertus Klei, Alexander Kolevzon, Itaru Kushima, Maria C. Lattig, So Lun Lee, Terho Lehtimäki, Lindsay Liang, Carla Lintas, Alicia Ljungdahl, Andrea del Pilar Lopez, Caterina Lo Rizzo, Yunin Ludena, Patricia Maciel, Behrang Mahjani, Nell Maltman, Marianna Manara, Dara S. Manoach, Dalia Marquez, Gal Meiri, Idan Menashe, Judith Miller, Nancy Minshew, Matthew Mosconi, Marina Natividad Avila, Rachel Nguyen, Norio Ozaki, Aarno Palotie, Mara Parellada, Maria Rita Passos-Bueno, Lisa Pavinato, Katherine P. Peña, Minshi Peng, Margaret Pericak-Vance, Antonio M. Persico, Isaac N. Pessah, Thariana Pichardo, Kaija Puura, Abraham Reichenberg, Alessandra Renieri, Kathryn Roeder, Catherine Sancimino, Stephan J. Sanders, Sven Sandin, F. Kyle Satterstrom, Stephen W. Scherer, Sabine Schlitt, Rebecca J. Schmidt, Lauren Schmitt, Katja Schneider-Momm, Paige M. Siper, Laura Sloofman, Moyra Smith, Renee Soufer, Christine R. Stevens, Pål Suren, James S. Sutcliffe, John A. Sweeney, Michael E. Talkowski, Flora Tassone, Karoline Teufel, Elisabetta Trabetti, Slavica Trajkova, Maria del Pilar Trelles, Brie Wamsley, Jaqueline Y. T. Wang, Lauren A. Weiss, Mullin H. C. Yu, Ryan Yuen, Jessica Zweifach.

## Introduction

Autism spectrum disorder (ASD) is characterized by deficits in social communication and the presence of restricted interests and/or repetitive behaviors (Lord *et al*., 2020). While the majority of the genetic liability for ASD is attributed to common genetic variation, rare variants, often arising *de novo*, play a substantial role in individual liability (Klei *et al*., 2012; Gaugler *et al*., 2014). Multiple large-scale studies of rare and common variation associated with ASD risk are ongoing, and dozens of new high-confidence risk genes have emerged (Fu *et al*., 2022; Zhou *et al*., 2022), primarily coding for proteins involved in gene expression regulation and neuronal communication or cytoskeleton (Satterstrom *et al*., 2020). Such findings have led to improvements in clinical care and serve as crucial, initial steps towards creating novel treatments and personalized interventions. Gene-targeted therapies for rare genetic disorders in ASD and associated neurodevelopmental disorders (NDDs) have emerged as a very dynamic area of study in academia and industry (Davidson *et al*., 2022). The overwhelming majority of participants in gene discovery studies are of European (EUR) ancestry, even though they comprise only 16% of the global population (Fatumo *et al*., 2022). This limited window into risk architecture across ancestries could exacerbate pre-existing disparities in diagnostics and service use for ASD (Martin *et al*., 2019). Indeed, recent studies have reported high rates of inconclusive results after genetic testing in non-European individuals, likely because of uncertainty in interpreting genomic variants (Abul-Husn *et al*., 2023; Venner *et al*., 2024).

We established the Genomics of Autism in Latin American Ancestries (GALA) Consortium to investigate the impact of genetic and environmental factors on ASD across Latin Americans – including participants from all of the Americas, corresponding to the Admixed American (AMR) superpopulation in the 1000 Genomes Project. Native Americans are thought to have originated in Northeast Asia (Hoffecker *et al*., 2023), with subsequent variation among existing Native American populations likely due to regional differentiation (Moreno-Mayar et al., 2018) and additional migration across the Bering Strait. The post-Columbian movement of people into the Americas added to this genetic diversity by admixture of Native American, African, and European populations. These AMR individuals comprise the largest recently-admixed population in the world, and the largest racial or ethnic minority in the United States. It is as yet unknown whether the genetic architecture of ASD differs across ancestral populations, and the genetic diversity of the AMR group (Moreno-Estrada *et al*., 2014; Ongaro *et al*., 2019) makes this question especially relevant.

We present the largest sequencing study to date of ASD in Latin American individuals and compare our results to findings from non-AMR cohorts. We show that a common measure of evolutionary impact on gene-level genetic variation, *i.e.*, genomic constraint scores, differ by ancestry. Yet, this is not the case for the most constrained genes, which are depauperate of population-level variation that is expected based on sequence composition of these genes. This is important because most, if not all, identified ASD-associated genes are evolutionarily constrained (Kosmicki *et al*., 2017; Fu *et al*., 2022), and this applies over diverse populations. Using Bayesian models, we identify 35 genome-wide significant ASD risk genes in Latin Americans, and observe a great degree of overlap with findings in largely European cohorts. We conclude that ASD and other NDD genes are shared across ancestry and that existing genetic testing pipelines are effective for the most deleterious variation, especially if information on allele frequency across ancestries is incorporated. We conclude that the biology of ASD is likely universal, and not impacted to any detectable degree by ancestry.

## Results

### Rare variant landscape in Latin Americans diagnosed with ASD

There are currently 10 GALA collection sites across the Americas, with data from seven sites (Brazil, California, Colombia, Costa Rica, Florida, New York, and Peru) included in this study (**Fig. 1**). Some GALA samples were sequenced as part of other large-scale whole exome sequencing (WES) and whole genome sequencing (WGS) efforts (Fu *et al*., 2022; DeFelice *et al*., 2024); analyses of 1,613 samples (including 707 ASD probands) are reported here for the first time. The GALA analyses reported here include all sequenced samples collected at GALA sites, as well as additional genetically-inferred AMR samples from SPARK and the Autism Sequencing Consortium (Buxbaum *et al*., 2012; SPARK Consortium., 2018).

**Figure 1.**
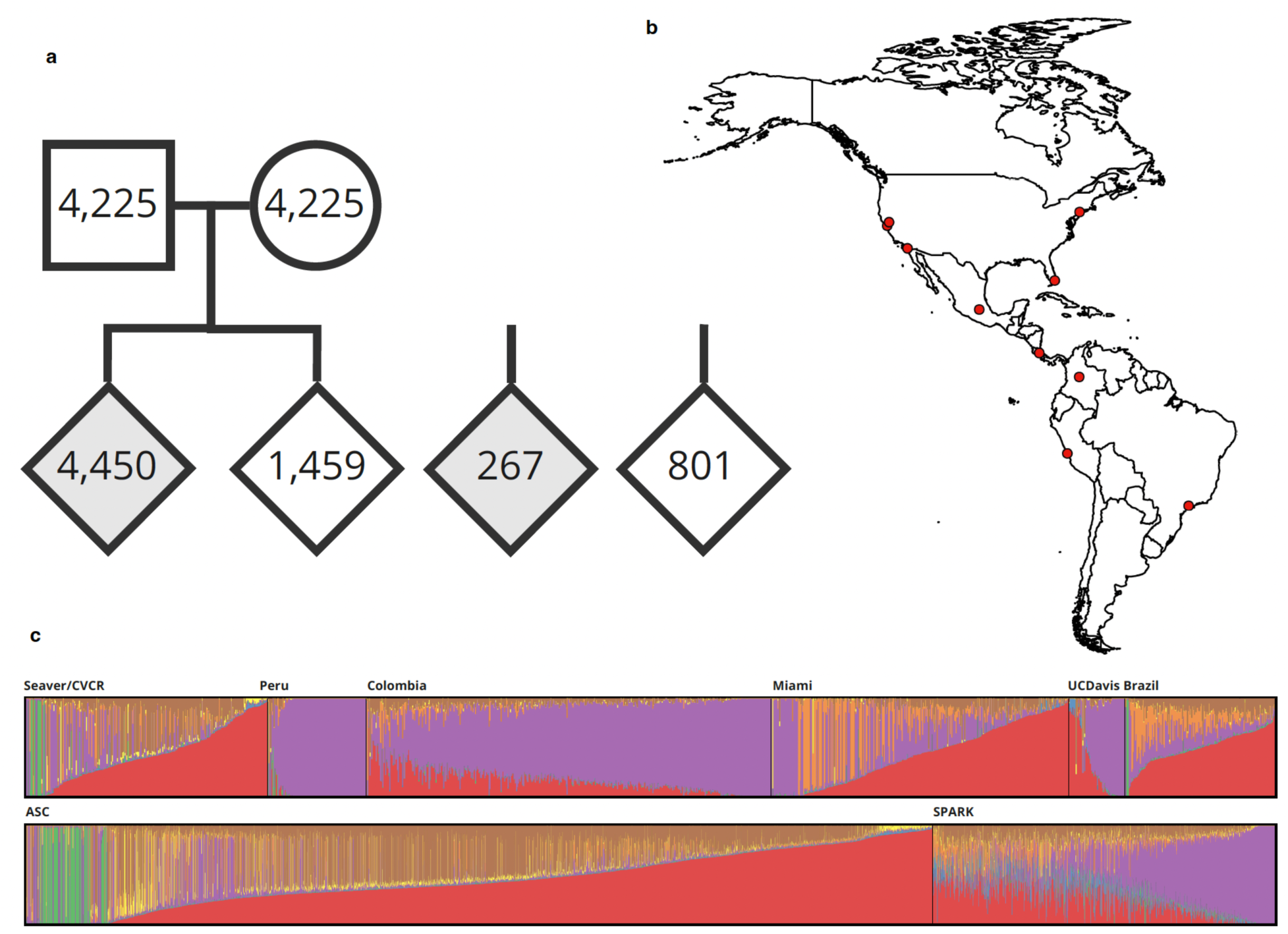
Overview of GALA cohort sites, structure, and ancestry composition. (a) Pedigree structure of the GALA cohort, comprising 4,717 cases and 10,710 controls. Diamonds represent offspring, probands are shown in gray, and typically-developing siblings in white. (b) Map of GALA collection sites across the Americas. (c) Ancestral composition of individuals from each GALA site based on ADMIXTURE analysis (K = 7). Ancestry components are: AMR (Admixed American, purple), NFE (non-Finnish European, red), AFR (African, orange), MID (Middle Eastern, brown), FIN (Finnish, blue), SAS (South Asian, yellow), and EAS (East Asian, green). AMR samples from the ASC and SPARK are presented in the lower panel.

A substantial source of individual ASD risk can reside in rare deleterious variation in conserved genes (Satterstrom *et al*., 2020; Fu *et al*., 2022), often *de novo* or very recent. Hence, to maximize power for discovery, we focus on data collected from family trios, *i.e*. an affected proband and both unaffected parents, and their typically-developing sibling(s) when available. When parental DNA samples could not be collected, we incorporated probands using a case-control framework. After extensive quality control (**Supplemental Fig. 1**; approaches adapted from (Satterstrom *et al*., 2020)), our dataset included *de novo* or case-control variants from 6,977 individuals, of which 4,717 had ASD (**Fig. 1b, Supplementary Table 1**). All together, 15,427 individuals, including parents when available, were sequenced with either WES (N = 14,152) or WGS (N = 207). Specifically, 14,359 individuals were sequenced as part of family-based analysis: 4,450 AMR ASD individuals (probands), 1,459 typically-developing siblings, and 8,450 parents. For case-control analysis, 267 AMR samples from individuals with ASD were matched to 801 non-psychiatric AMR controls from the Mount Sinai BioMe biobank (Abul-Husn *et al*., 2021; Belbin *et al*., 2021).

We identified 6,555 rare [*i.e.*, allele frequency < 0.1% in our dataset and in the population-specific non-neuro subsets of gnomAD v2.1.1 and gnomAD v3.1.2 (Karczewski *et al*., 2020; Chen *et al*., 2023)], unique *de novo* single nucleotide variants (SNVs) or insertion deletion variants (indels) (5,062 in ASD probands and 1,493 in unaffected siblings) in protein-coding exons (**Supplementary Table 2**). Additionally, we identified 36 variants that occurred twice, with 18 being unique to particular families, consistent with germline mosaicism. Finally, we also observed 211 and 15 rare autosomal *de novo* small genic CNVs in 2,191 probands and 707 siblings, respectively (**Supplementary Table 3**).

In prior studies, highly constrained genes showed an aggregated signal of ASD risk-conferring variants (Kosmicki *et al*., 2017) and integrating genomic constraint scores has proven powerful for gene discovery (Fu *et al*., 2022). However, genomic constraint scores are derived from cohorts largely of European ancestry. Therefore, we first sought to evaluate the utility of these scores on samples of diverse ancestry.

First, we examined the distribution of *de novo* variants as a function of a well-established metric of tolerance to loss-of-function variants, the loss-of-function observed/expected upper bound fraction (LOEUF) (Karczewski *et al*., 2020). Genes with low LOEUF scores are depleted for loss-of-function variation as result of negative natural selection (Karczewski *et al*., 2020). Our results demonstrate that rates of *de novo* variants for both protein truncating (PTV) and deleterious missense (MisB, with a ‘missense badness, PolyPhen-2, and constraint’ (MPC) score ≥ 2 (Samocha *et al*., 2017)) variants are elevated in probands compared to unaffected siblings in genes with low LOEUF scores (**Fig. 2**). Comparing our findings with previously published results (Fu *et al*., 2022), we observed that the rates of *de novo* variation are consistent when comparing all AMR to all other samples (**Supplementary Fig. 2**).

**Figure 2.**
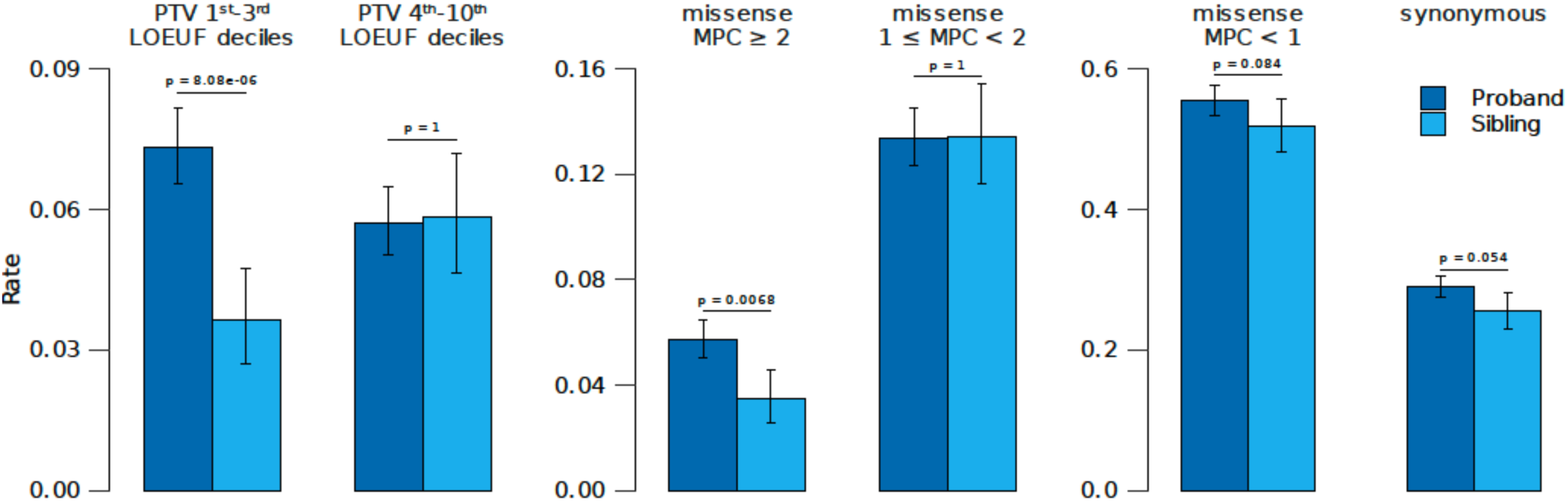
Comparison of rare de novo variant counts per sample between ASD probands and unaffected siblings in the GALA cohort. The average number of rare variants per sample is compared between ASD probands (dark blue, n = 4,450) and unaffected siblings (light blue, n = 1,459) of Admixed American ancestry (AMR). The analysis includes (**left**) protein truncating variants (PTVs) in highly constrained genes (LOEUF deciles 1–3, 5,363 genes) and less constrained genes (LOEUF deciles 4–10, 12,765 genes), (**middle**) missense variants categorized by predicted functional severity (MPC ≥ 2 for high severity, 1 ≤ MPC < 2 for moderate severity), and (**right**) MPC < 1 (for low severity) and synonymous missense variants. Error bars represent 95% confidence intervals.

Second, we examined whether LOEUF is well calibrated across ancestral populations. Effective population size differs across Native American, European and African populations (Browning *et al*., 2018), but current estimates of gene constraint are derived from cohorts that are largely of European ancestry. Existing LOEUF scores are modestly over-conservative when applied to AMR samples (**Fig. 3, Supplemental Fig. 3** and (Karczewski *et al*., 2020)), but, when focusing on the most constrained (lower) deciles, correlate well with the observed number of PTVs normalized by sample size and gene length (**Fig. 3**). Since association signal concentrates to these lower deciles (**Fig. 2**), these observations justify the use of existing LOEUF scores for our study, and generally for studies focusing on highly constrained genes in other ancestries, including admixed African ancestries (**Supplemental Fig. 3**).

**Figure 3.**
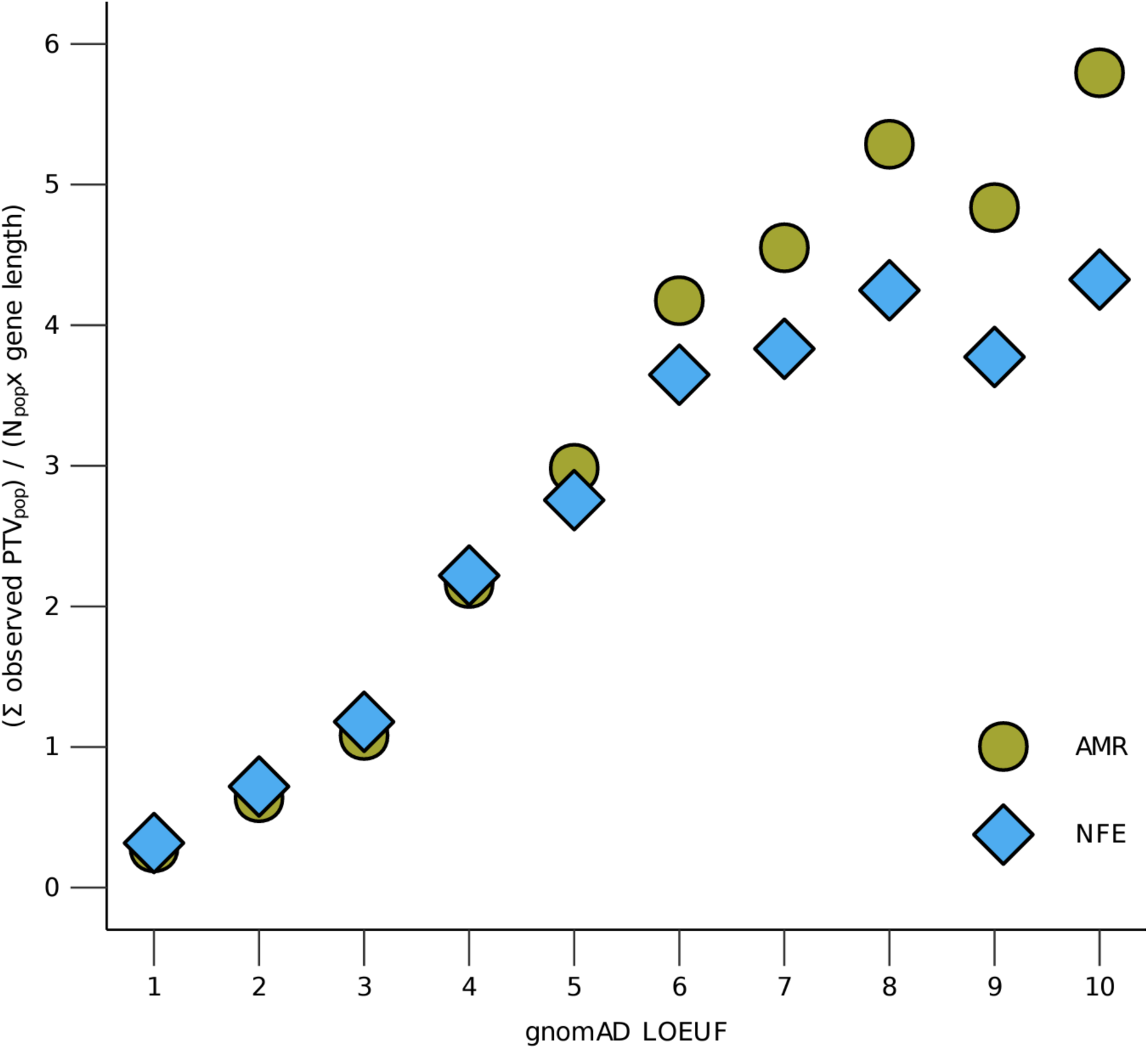
Ratio of observed PTVs in NFE vs. AMR ancestries. The sum of observed protein truncating variants (PTVs) is plotted for Non-Finnish European (NFE) and Admixed American (AMR) populations in gnomAD v2.1.1, scaled to population size and total coding sequence length for each gnomAD LOEUF decile. Population sizes are NFE: 56,885 and AMR: 17,296. LOEUF deciles reflect gene constraint, with lower deciles indicating more constrained genes.

### ASD Gene Discovery in Latin Americans

For gene discovery, we used TADA (Transmission And *De novo* Association), an algorithm developed to integrate *de novo*, inherited, and case-control variants, as well as, more recently, LOEUF scores and small genic CNVs (He *et al*., 2013; De Rubeis *et al*., 2014; Satterstrom *et al*., 2020; Fu *et al*., 2022). Sixteen genes were associated with ASD at FDR < 0.01, 35 meeting genome-wide significant association (FDR < 0.05), and 61 genes associated at FDR < 0.1 (**Fig. 4**; **Table 1; Supplementary Table 4**). To formally examine the overlap of these findings with those in largely EUR ASD cohorts, we first identified and removed all AMR samples in Fu *et al*. (Fu *et al*., 2022), yielding a non-AMR complementary set (Fu_COMP_) with no overlap with our analyses. Nineteen of the 35 GALA genes with FDR < 0.05 showed significant signal in Fu_COMP_. We next compared the observed numbers of variants in the GALA cohort with the expected number of variants derived from TADA analysis in Fu_COMP_. To do this, we compared results for concordant genes, defined as genes that show FDR < 0.05 in GALA and in Fu *et al. (Fu et al., 2022)*; when we compare the numbers of variants in the GALA cohort with the expected number of variants derived from TADA analysis in Fu_COMP_ the findings are consistent with expectation (**Supplemental Material, Table 2**). We also compared our gene findings from the GALA cohort with those in a large cohort ascertained for severe developmental disorders (DDs) (Kaplanis *et al*., 2020): Six of the 16 genes that had an FDR < 0.05 in GALA and an FDR > 0.1 in Fu_COMP_, showed an FDR < 0.05 in the DD cohort (**Table 1**). Furthermore, amongst the 35 GALA ASD genes with FDR < 0.05, 26 are noted as Strong or Definitive DD genes in DECIPHER (Firth *et al*., 2009), and 29 have an OMIM Morbid Map (McKusick, 1998) designation with a neurological or psychiatric phenotype.

**Figure 4.**
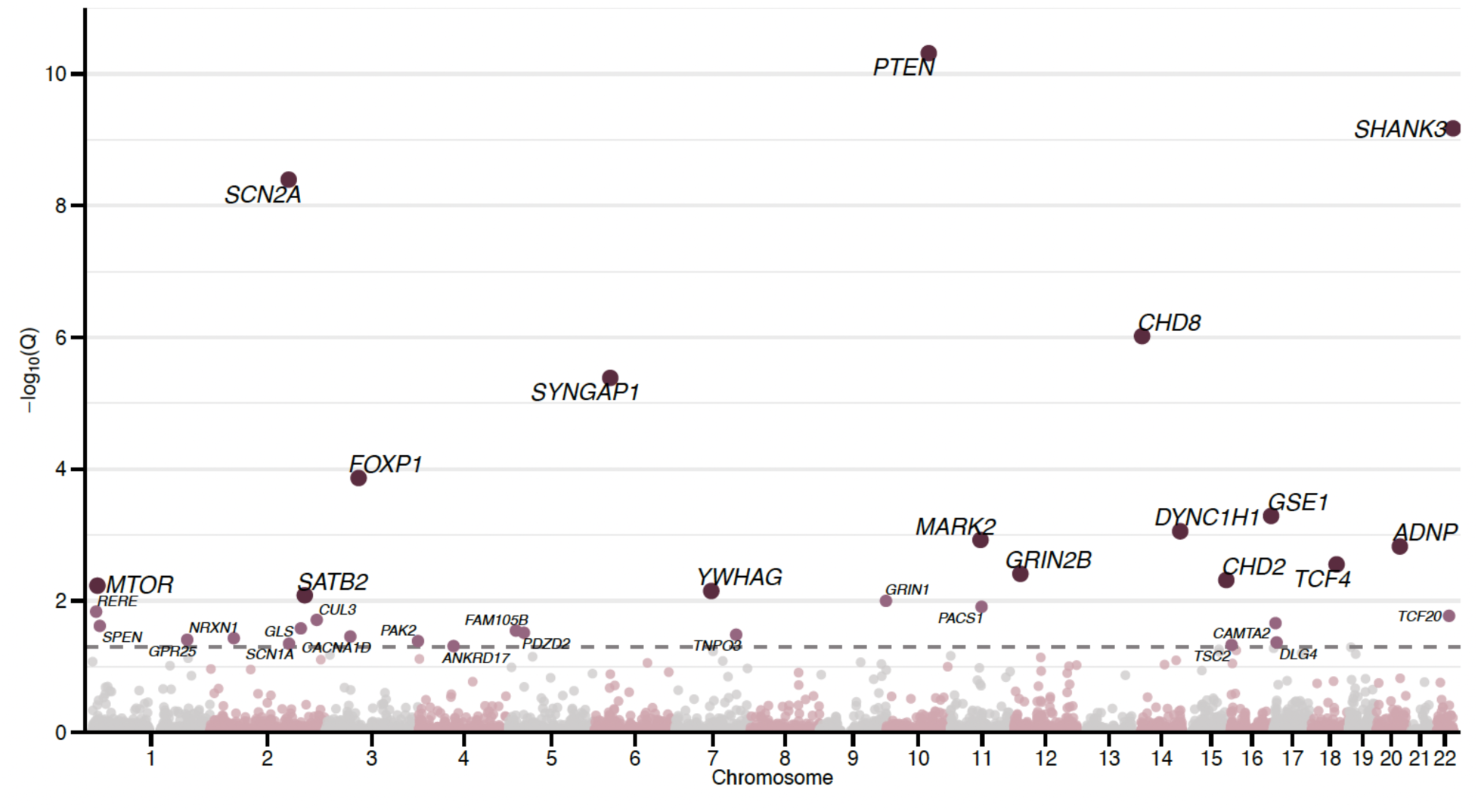
Manhattan plot of ASD genes identified in Latin American participants. The plot displays 35 genes identified with a false discovery rate (FDR) threshold of < 0.05 (see **Table 1**).

**Table 1.**
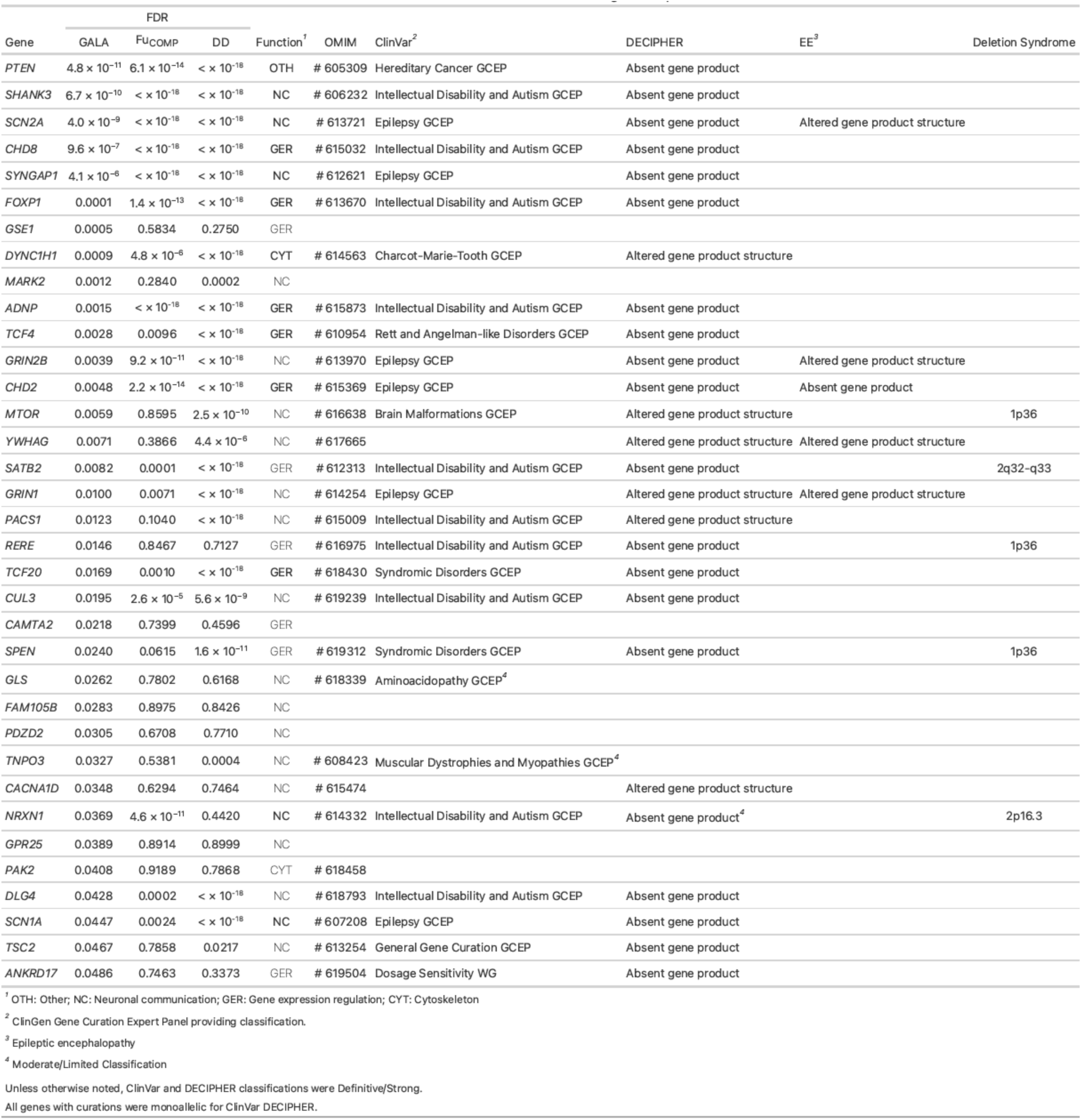
Genome-wide and clinical findings for top 35 genes. For genes with FDR < 0.05 in GALA, we show the FDR in Fu_COMP_ and in DD. In addition, we annotated the genes for function, as was done in Satterstrom et al. We retained annotations from Satterstrom (bold) and annotated the remaining genes in a similar manner. For each gene, we also reviewed OMIM Morbid Map, ClinVar and the DECIPHER v11.28 website. All genes that have a CNS or PNS OMIM Morbid designation are noted. For ClinVar all genes that have Strong or Definitive autosomal dominant findings are noted, together with the Gene Curation Expert Panel that made the designation. Two genes with Moderate or Limited evidence are noted. Similarly, all genes that show a Strong or Definitive autosomal dominant finding in DECIPHER are noted, along with the proposed mechanism. If an epileptic encephalopathy was also noted, then the proposed mutation mechanism was noted for that as well. Note again that one gene (*NRXN1*) had only Moderate evidence in DECIPHER.

| Gene | FDR |  |  | Function<sup>1</sup> | OMIM | ClinVar<sup>2</sup> | DECIPHER | EE<sup>3</sup> | Deletion Syndrome |
| --- | --- | --- | --- | --- | --- | --- | --- | --- | --- |
|  | GALA | F<sub>U</sub>COMP | DD |  |  |  |  |  |  |
| *PTEN* | 4.8 × 10<sup>-11</sup> | 6.1 × 10<sup>-14</sup> | < × 10<sup>-18</sup> | OTH | # 605309 | Hereditary Cancer GCEP | Absent gene product |  |  |
| *SHANK3* | 6.7 × 10<sup>-10</sup> | < × 10<sup>-18</sup> | < × 10<sup>-18</sup> | NC | # 606232 | Intellectual Disability and Autism GCEP | Absent gene product |  |  |
| *SCN2A* | 4.0 × 10<sup>-9</sup> | < × 10<sup>-18</sup> | < × 10<sup>-18</sup> | NC | # 613721 | Epilepsy GCEP | Absent gene product | Altered gene product structure |  |
| *CHD8* | 9.6 × 10<sup>-7</sup> | < × 10<sup>-18</sup> | < × 10<sup>-18</sup> | GER | # 615032 | Intellectual Disability and Autism GCEP | Absent gene product |  |  |
| *SYNGAP1* | 4.1 × 10<sup>-6</sup> | < × 10<sup>-18</sup> | < × 10<sup>-18</sup> | NC | # 612621 | Epilepsy GCEP | Absent gene product |  |  |
| *FOXP1* | 0.0001 | 1.4 × 10<sup>-13</sup> | < × 10<sup>-18</sup> | GER | # 613670 | Intellectual Disability and Autism GCEP | Absent gene product |  |  |
| *GSE1* | 0.0005 | 0.5834 | 0.2750 | GER |  |  |  |  |  |
| *DYNC1H1* | 0.0009 | 4.8 × 10<sup>-6</sup> | < × 10<sup>-18</sup> | CYT | # 614563 | Charcot-Marie-Tooth GCEP | Altered gene product structure |  |  |
| *MARK2* | 0.0012 | 0.2840 | 0.0002 | NC |  |  |  |  |  |
| *ADNP* | 0.0015 | < × 10<sup>-18</sup> | < × 10<sup>-18</sup> | GER | # 615873 | Intellectual Disability and Autism GCEP | Absent gene product |  |  |
| *TCF4* | 0.0028 | 0.0096 | < × 10<sup>-18</sup> | GER | # 610954 | Rett and Angelman-like Disorders GCEP | Absent gene product |  |  |
| *GRIN2B* | 0.0039 | 9.2 × 10<sup>-11</sup> | < × 10<sup>-18</sup> | NC | # 613970 | Epilepsy GCEP | Absent gene product | Altered gene product structure |  |
| *CHD2* | 0.0048 | 2.2 × 10<sup>-14</sup> | < × 10<sup>-18</sup> | GER | # 615369 | Epilepsy GCEP | Absent gene product | Absent gene product |  |
| *MTOR* | 0.0059 | 0.8595 | 2.5 × 10<sup>-10</sup> | NC | # 616638 | Brain Malformations GCEP | Altered gene product structure |  | 1p36 |
| *YWHAG* | 0.0071 | 0.3866 | 4.4 × 10<sup>-6</sup> | NC | # 617665 |  | Altered gene product structure | Altered gene product structure |  |
| *SATB2* | 0.0082 | 0.0001 | < × 10<sup>-18</sup> | GER | # 612313 | Intellectual Disability and Autism GCEP | Absent gene product |  | 2q32-q33 |
| *GRIN1* | 0.0100 | 0.0071 | < × 10<sup>-18</sup> | NC | # 614254 | Epilepsy GCEP | Altered gene product structure | Altered gene product structure |  |
| *PACS1* | 0.0123 | 0.1040 | < × 10<sup>-18</sup> | NC | # 615009 | Intellectual Disability and Autism GCEP | Altered gene product structure |  |  |
| *RERE* | 0.0146 | 0.8467 | 0.7127 | GER | # 616975 | Intellectual Disability and Autism GCEP | Absent gene product |  | 1p36 |
| *TCF20* | 0.0169 | 0.0010 | < × 10<sup>-18</sup> | GER | # 618430 | Syndromic Disorders GCEP | Absent gene product |  |  |
| *CUL3* | 0.0195 | 2.6 × 10<sup>-5</sup> | 5.6 × 10<sup>-9</sup> | NC | # 619239 | Intellectual Disability and Autism GCEP | Absent gene product |  |  |
| *CAMTA2* | 0.0218 | 0.7399 | 0.4596 | GER |  |  |  |  |  |
| *SPEN* | 0.0240 | 0.0615 | 1.6 × 10<sup>-11</sup> | GER | # 619312 | Syndromic Disorders GCEP | Absent gene product |  | 1p36 |
| *GLS* | 0.0262 | 0.7802 | 0.6168 | NC | # 618339 | Aminoacidopathy GCEP<sup>4</sup> |  |  |  |
| *FAM105B* | 0.0283 | 0.8975 | 0.8426 | NC |  |  |  |  |  |
| *PDZD2* | 0.0305 | 0.6708 | 0.7710 | NC |  |  |  |  |  |
| *TNPO3* | 0.0327 | 0.5381 | 0.0004 | NC | # 608423 | Muscular Dystrophies and Myopathies GCEP<sup>4</sup> |  |  |  |
| *CACNA1D* | 0.0348 | 0.6294 | 0.7464 | NC | # 615474 |  | Altered gene product structure |  |  |
| *NRXN1* | 0.0369 | 4.6 × 10<sup>-11</sup> | 0.4420 | NC | # 614332 | Intellectual Disability and Autism GCEP | Absent gene product<sup>4</sup> |  | 2p16.3 |
| *GPR25* | 0.0389 | 0.8914 | 0.8999 | NC |  |  |  |  |  |
| *PAK2* | 0.0408 | 0.9189 | 0.7868 | CYT | # 618458 |  |  |  |  |
| *DLG4* | 0.0428 | 0.0002 | < × 10<sup>-18</sup> | NC | # 618793 | Intellectual Disability and Autism GCEP | Absent gene product |  |  |
| *SCN1A* | 0.0447 | 0.0024 | < × 10<sup>-18</sup> | NC | # 607208 | Epilepsy GCEP | Absent gene product |  |  |
| *TSC2* | 0.0467 | 0.7858 | 0.0217 | NC | # 613254 | General Gene Curation GCEP | Absent gene product |  |  |
| *ANKRD17* | 0.0486 | 0.7463 | 0.3373 | GER | # 619504 | Dosage Sensitivity WG | Absent gene product |  |  |

<sup>1</sup> OTH: Other; NC: Neuronal communication; GER: Gene expression regulation; CYT: Cytoskeleton
<sup>2</sup> ClinGen Gene Curation Expert Panel providing classification.
<sup>3</sup> Epileptic encephalopathy
<sup>4</sup> Moderate/Limited Classification
Unless otherwise noted, ClinVar and DECIPHER classifications were Definitive/Strong.
All genes with curations were monoallelic for ClinVar DECIPHER.

As in prior studies, *de novo* variation provided a major source of signal for top genes (**Supplemental Fig. 4**). Similarly, PTVs are a major source of variation, and it was interesting to note that missense variants, and particularly MisB variants, were also an important source of rare variation association signal (**Supplemental Fig. 5**). For several of the top genes, the association signal is fully or almost fully derived from missense variants in the GALA cohort, which for *MTOR*, *YWHAG*, *GRIN1*, *PACS1*, and *CACNA1D*, is consistent with prior findings (“Altered gene product structure” in DECIPHER; see **Table 1**).

### Implications for clinical genetics

With compelling evidence for overlapping ASD gene findings in AMR samples, we next asked about the fraction of findings that are potentially reportable as per the American College of Medical Genetics (ACMG) guidelines for sequence variants (Richards *et al*., 2015). We used the VarSome package (Kopanos *et al*., 2018) — minimizing the use of proprietary databases and approaches used by commercial testing laboratories — to evaluate (1) genome-wide *de novo* variation and (2) inherited variation in a list of well-defined X-linked risk genes (**Supplementary Table 5**). We analyzed all GALA samples and all Fu_COMP_ samples, focusing on genes for which there was a reported association with an ASD phenotype and/or a broader NDD phenotype (see Methods).

In our *de novo* analysis, we focused on 20,571 variants. Among these, 926 (4.5%) were classified by VarSome as pathogenic or likely pathogenic (P/LP) when we focused on genes that included ASD among the associated phenotypes (**Supplementary Table 6**). In the AMR cohort (N = 4,450), 195 *de novo* variants (3.76%, 95% CI 3.27-4.32%) were identified as P/LP (**Supplementary Table 8**), compared to 731 out of 15,386 *de novo* variants (4.75%, 95% CI 4.42-5.10%) in non-AMR samples. In terms of participants with findings, 4.31% (95% CI 3.75-4.96%) of AMR probands and 5.53% (95% CI 5.15-5.94%) of non-AMR probands had at least one P/LP variant identified. Comparisons between EUR and non-EUR participants revealed that EUR individuals had a higher rate of *de novo* P/PL variants. Specifically, EUR participants had 634 (4.83%, 95% CI 4.47-5.21%) *de novo* P/LP variants identified, compared to 292 (3.92%, 95% CI 3.50-4.40%) in non-EUR participants. Overall, EUR participants had a higher rate of P/LP variants identified, as compared to non-EUR participants (5.61%, 95% CI 5.20-6.06% vs 4.54%, 95% CI 4.05-5.09%).

When broadening our criteria from ASD to include other known NDD phenotypes, 1,339 *de novo* variants were deemed to be P/LP (**Supplementary Table 7**). In AMR 276 *de novo* variants were classified as P/LP (5.32%, 95% CI 4.74-5.98%), versus 1,063 in non-AMR individuals (6.91%, 95% CI 6.52-7.32%). In terms of participants with *de novo* findings, 6.07% (95% CI 5.39-6.82%) of AMR participants and 7.99% (95% CI 7.53-8.47%) of non-AMR participants had at least one likely P/LP finding. EUR participants had a notably higher rate of findings (8.22%, 95% CI 7.72-8.75%) compared to 6.24% (95% CI 5.66-6.87%) in non-EUR individuals.

Extending our analysis to include X-linked inherited findings, we observed a further increase in P/LP detection rates. Specifically, 201 *de novo* or X-linked variants (4.67%, 95% CI 4.24-5.13%) in AMR samples and 758 variants (3.58%, 95% CI 4.24-5.13%) in non-AMR samples were classified as P/LP for ASD. When we broadened the terms to include other NDD-related genes, the proportion of P/LP variants rose to 4.90% (95% CI 4.36-5.58%) in AMR individuals and 5.26% (95% CI 4.96-5.57%) in non-AMR participants. The rate of participants with at least one P/LP variant increased to 6.47% (95% CI 5.78-7.24%) in AMR samples and 8.38% (95% CI 7.91-8.87%) of non-AMR participants (**Supplementary Table 8**). Comparing EUR and non-EUR groups, EUR participants showed a higher yield of P/LP findings (8.62%, 95% CI 8.11-9.16%) compared to 6.63% (95% CI 6.04-7.28%) of non-EUR individuals.

Qualitatively similar results were made when using Neptune (Eric *et al*., 2021), which uses databases of previously identified variants to call P/LP in a set of target genes (**Supplemental Fig. 6 and Supplemental Materials**). There are more rare variants identified in diverse samples together with a lower rate of classification. These two opposing findings suggest a reduction in findings per individual in AMR or non-EUR individuals, when compared to non-AMR or EUR, respectively. Considering the VarSome and Neptune results together, the findings provide support for the translatability of rare genetic findings in ASD across ancestries in a clinical setting, albeit with opportunities for improvement.

## Discussion

The past decade has seen significant advances in deciphering the genetic architecture of ASD, but largely from EUR cohorts. It is not yet known whether the genetic architecture of ASD differs across ancestral populations, including in admixed populations. Latin American individuals comprise the largest recently-admixed population in the world, and the largest racial or ethnic minority in the United States. Diverse sites with large AMR representation have joined to form GALA, and here we report a first, large-scale multinational analysis of rare variant risk in Latin Americans with ASD, identifying ASD-associated genes in this cohort and comparing genetic architecture with that observed in non-AMR ASD.

As in previous studies, we found that signal for ASD risk genes was concentrated in highly conserved genes and largely driven by very rare de novo variation. For the discovery of ASD genes impacted by very rare de novo or case-control variation, it is critical to have reliable estimates of expected genic mutation rates, which can be derived from both cross-species comparisons, and empirical data from massive aggregated sequencing resources, such as gnomAD. While representation of diverse populations is improving, much of the existing sequence data are skewed towards EUR samples. Thus, there is much more to be done regarding genetic variability within under-represented populations. Our analyses confirm that metrics of gene-level constraint are overly conservative, due to the over-reliance on EUR samples that have a lower effective population size. However, we also demonstrate that the key metric LOEUF — when applied to the most conserved genes — is well calibrated across diverse ancestral populations.

Since deleterious variation in highly conserved genes is subject to strong purifying selection, such variation is both very rare and frequently *de novo*. Allele frequency filtering based on gnomAD or other such datasets is hence an important means to infer very rare variation. However, we observe that relying on overall allele frequency allows for the introduction of more common variation into the analyses, hence reducing power and increasing the false positive rate. We began our analyses using established best practices for filtering by global allele frequency in the analysis of potentially *de novo* variants (Satterstrom *et al*., 2020; Fu *et al*., 2022). However, we noticed that some variants initially classified as rare in gnomAD (AF < 0.1%) turned out to be more common in particular populations. For example, PTV 10:32491945:C:T in *CCDC7* (a gene with a LOEUF score of 0.982) has an overall gnomAD frequency of 0.053%, but 0.362% in the AMR subpopulation, while PTV 2:38825971:G:A in *DHX57* (LOEUF score of 0.675) has an overall gnomAD frequency of 0.0048% but of 0.129% in the Ashkenazi Jewish subpopulation. To address this heterogeneity, we recommend annotating variants with allele frequencies across all subpopulations in the non-neuro releases of gnomAD versions 2.1.1 and 3.1.2., as we have done here. Building on this strategy, we extended the same annotation to our analysis of inherited variation, adopting a more stringent allele frequency threshold of < 0.01%, to ensure even more precision in our findings (Arriaga-MacKenzie *et al*., 2021; Gudmundsson *et al*., 2022).

We next used TADA to associate 35 genes with ASD at an FDR threshold < 0.05 from the GALA data, 16 with FDR < 0.01, and 8 with FDR < 0.001 (**Fig. 4**; **Table 1**). Consistent with prior studies in largely EUR cohorts, gene expression regulation, neuronal communication, and cytoplasmic genes are well represented in the GALA ASD genes (**Table 1 and Supplemental Tables 17and 18**). FDR is well calibrated in TADA (Satterstrom *et al*., 2020), and genes identified with TADA in smaller cohorts are consistently replicated at expected levels in larger samples. However, it is still important to evaluate the level of confidence in the genes identified. First, as noted above, we compared results for top genes across GALA and a recent large-scale study (FDR < 0.05 in both AMR samples and non-AMR/Fu_COMP_ studies), and observed that findings are consistent with expectation (**Table 2**). However, there are multiple genes with evidence in GALA but not in Fu_COMP_. This can be for one of several reasons, including (1) sparseness of *de novo* events and, hence, over/under-representation of *de novo* events in subsamples, (2) differences in ascertainment, and (3) the possibility that some findings are false positive findings. While all three could make some contribution, (1) was extensively evaluated previously (Satterstrom *et al*., 2020; Fu *et al*., 2022) by simulation and other analyses, suggesting that (1) is likely to be the major contributor to discordance. This perspective receives support when we compared our gene findings from the GALA cohort with those in a large cohort ascertained for severe DDs (Kaplanis *et al*., 2020): Six of the 16 genes that had an FDR < 0.05 in GALA and an FDR > 0.1 in Fu_COMP_, showed an FDR < 0.05 in the DD cohort (**Table 1**). In addition, amongst the 35 ASD genes with an FDR < 0.05, most have a dominant neurodevelopmental morbid association in OMIM, DECIPHER, and/or ClinVar. The concordance of findings between genome-wide studies (GALA, Fu_COMP_ and DD) and curated clinical databases indicate that our approach is valid for ASD gene discovery in AMR samples, and that the false discovery rates are likely well calibrated.

We next examined specific emerging genes found in the GALA analyses, including contrasting results with those seen in non-AMR samples (Fu_COMP_) and curated databases. *MARK2* showed an FDR < 0.002 in GALA. It was not significant in FuCOMP and was not noted in DECIPHER or OMIM, but it showed signal in severe DD (**Table 1**). MARK2 (microtubule affinity-regulating kinase 2) is a serine/threonine kinase that plays a key role in phosphorylation of microtubule associated proteins and regulates the MTOR pathway (Lei, Zhang and Cai, 2024). *MARK2* is constrained against both PTV and missense variation, and has recently been implicated in ASD (Trost *et al*., 2022; Zhou *et al*., 2022; Gong *et al*., 2024). Results from the GALA cohort provide genome-wide support for *MARK2* as an ASD-associated gene (**Fig. 7**).

*MTOR*, *YWHAG*, *PACS1*, *TNPO3*, and *TSC2* did not show appreciable signal in Fu_COMP_, but all show signal in severe DD, have OMIM Morbid Map information, and – except for *TNPO3* – are flagged in DECIPHER. *SPEN* shows a trending signal in Fu_COMP_, a strong signal in DD, and is annotated in both DECIPHER and OMIM. *RERE*, *GLS*, *CACNA1D*, and *ANKRD17* are noted in both OMIM Morbid Map and DECIPHER, while *PAK2* has an OMIM Morbid Map designation.

Recurrent R230W mutations in PACS1 are responsible for PACS1 Neurodevelopmental Disorder (PACS1-NDD), also called Schuurs−Hoeijmakers syndrome (Arnedo et al., 2022) and mutations appear to have a direct impact on neuronal activity via impact on HDAC6 (Villar-Pazos et al., 2023; Rylaarsdam et al., 2024). GALA included two R230W mutations, but also a de novo R709Q mutation, which is potentially contributing to the ASD phenotype. We note that DECIPHER identifies multiple de novo variants at R230W in PACS1, but also identifies other de novo changes (**Fig. 5**).

**Figure 5.**
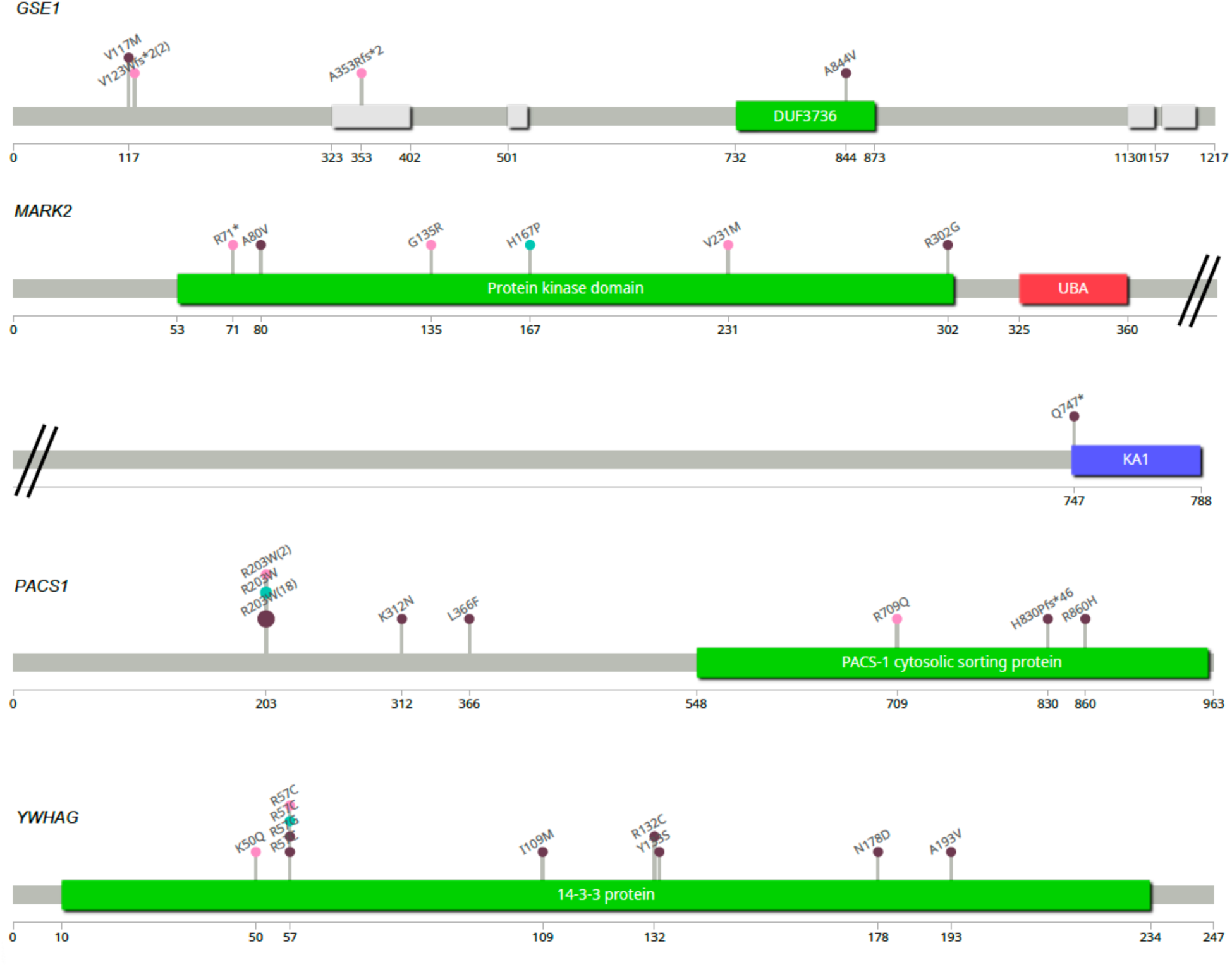
“Lollipop” diagrams illustrating variants identified in ASD-associated genes. Variants observed in GALA analyses of AMR individuals are marked with pink circles, those found in Fu_COMP_ individuals are marked with green, and variants found in DECIPHER are in purple. Note that there were two instances of V123Wfs*2 variants in *GSE1* in GALA, two instances of R203W in *PACS1* in GALA, and 18 such variants in DECIPHER. Figures were generated using the ‘lollipop’ software package (Jay and Brouwer, 2016).

Haploinsufficiency of *RERE*, *SPEN*, and *MTOR* are all thought to contribute to the neurodevelopmental phenotypes in proximal 1p36 deletion syndrome (**Table 1**) (Jacquin *et al*., 2023). *RERE* (arginine-glutamic acid dipeptide repeats) is a member of the Atrophin family of transcriptional repressors (Jordan *et al*., 2018); and, *SPEN*, also called SHARP (SMRT and HDAC-associated repressor protein), interacts with HDAC1 and HDAC2 and is involved in X-chromosome silencing (Radio *et al*., 2021). Heterozygous variants in *RERE* have been shown to cause neurodevelopmental disorder with or without anomalies of the brain, eye, or heart (NEDBEH; OMIM #616975). There is emerging evidence of possible genotype-phenotype correlations in *RERE*, with missense mutation in the Atrophin-1 domain associated with more severe, syndromal presentations (Jordan *et al*., 2018) and PTVs in this domain and elsewhere in the gene show a less severe phenotype. Two GALA participants carried PTVs in *RERE*, consistent with these findings (**Supplemental Fig. 7**). Mutations in *SPEN* lead to a neurodevelopmental disorder through heterozygous loss-of-function mutations (Radio *et al*., 2021), and we identified two *de novo* missense variants in this gene.

MTOR signaling is a prominent pathway amongst the top GALA genes: *PTEN* is a top finding, and *MTOR* and *TSC2* are identified as well. *CUL3* is a core component of the E3 ubiquitin ligase complex and indirectly regulates MTOR signaling (Chen *et al*., 2018). As noted above, MARK2 also regulates the MTOR pathway (Lei, Zhang and Cai, 2024). *GLS* is interesting in this context, because the genes codes for glutaminase and levels of glutamine and glutamate regulate the MTOR pathway (Csibi *et al*., 2013; Tan, Sim and Long, 2017; Takahara *et al*., 2020; Bodineau *et al*., 2021). There are recessive disorders associated with biallelic mutations in this gene (OMIM #618339 and #618328), and there is emerging evidence for a dominant neurodevelopmental disorder (OMIM #618339), as well as elevated risk for chronic kidney disease and liver disease associated with a DNA repeat expansion (Hujoel *et al*., 2024). We identified two *de novo* missense mutations (M465I in the glutaminase domain, and E596Q in the Ankyrin repeat domain) providing support for the heterozygous disorder; study of glutamine and glutamate in these participants will be of interest.

GLS also plays a more direct role in neuronal signaling in the brain, and the presence of *GLS*, *GRIN1*, *GRIN2B*, *SYNGAP1*, *DLG4*, *SHANK3*, *NRXN1*, *SCN2A*, *SCN1A*, and *CACNA1D* among the top genes is consistent with a large body of evidence implicating glutamatergic synaptic organization, plasticity and signaling in ASD. Two of the *CACNA1D* variants in our dataset (G1169V and A769G) are also deemed pathogenic using a recently described CACNA1D-specific model (Tang *et al*., 2024). The 14-3-3 family of proteins are scaffolding proteins that integrate signals in neurons and other cells (Pitasse-Santos *et al*., 2024). For example, 14-3-3ɛ is a PPI Hub Protein that interacts with several top GALA genes (i.e., *DLG4*, *TSC2*, *GRIN2B*, *MARK2*, *GRIN1*, *YWHAG) using Enrichr (Xie et al., 2021)*. *YWHAG* codes for 14-3-3ɛ, and is significant in GALA and in the DD cohort and has a neurodevelopmental finding in DECIPHER. Missense mutations in Arg 57 were observed in GALA, Fu_COMP_, and in DECIPHER, and a nearby missense mutation in Lys 50 was observed in GALA (**Fig. 5**).

In addition to *MARK2*, *DYNC1H1* and *PAK2* further implicate cytoskeletal organization and intracellular transport in ASD. *DYNC1H1* codes for cytoplasmic dynein heavy chain, which forms the core of cytoplasmic dynein – the main retrograde motor in cells (Dahl *et al*., 2021). Early and later onset neuromuscular and developmental disorders have been associated with missense and PTV mutation in *DYNC1H1* (Möller *et al*., 2024). PAK2 is a serine/threonine kinase that activates the LIMK-cofilin pathway, a key regulator of synaptic actin dynamics (Zhang *et al*., 2022), and we observed a *de novo* frameshift (P30Lfs*8) and missense (M436Y) mutation in *PAK2*, the latter in the protein kinase domain. Although *PAK2* is not annotated for DD and DECIPHER and is not yet reviewed in ClinVar, we consider that it is a significant ASD-associated gene.

*TNPO3* is interesting since both OMIM and ClinGen consider it a gene for a limb-girdle muscular dystrophy (Costa *et al*., 2022). Across GALA and Fu_COMP_ there were five *de novo* missense mutations throughout the molecule, which is in contrast to the COOH-terminal mutations described in the limb-girdle muscular dystrophy syndrome (**Supplemental Fig. 7**) (Costa *et al*., 2022). While mutations in *DMD* can lead to ASD (Geuens *et al*., 2024), further genotype-phenotype studies are indicated to understand whether different mutations in *TNPO3* have psychiatric and/or muscular phenotypes.

Heterozygous mutations in *ANKRD17* are the cause of Chopra-Amiel-Gordon syndrome (CAGS; OMIM #619504), which is a neurodevelopmental disorder characterized by variable levels of global developmental delay and/or impaired intellectual development. ASD has been observed in some cases, and our results in GALA and in Fu_COMP_ (**Supplemental Fig. 7**) support this gene as an ASD-associated gene.

Finally, we consider *GSE1* as a potential ASD-associated gene based on findings in GALA (**Fig. 5**). *GSE1* was not supported by other studies noted in Table 1, however, an examination of the newest (and largest) ASC dataset this gene shows genome-wide significance (FDR=0.00091) with five *de novo* PTVs in cases (A353Rfs*2, V123Wfs*2(2), G36Afs*8 and Q340*) and none in unaffected siblings (Satterstrom *et al*., 2024). GSE1, which complexes with the HDAC1/CoREST complex (Vcelkova *et al*., 2023) provides further evidence for gene expression regulation in ASD.

Altogether, the results are consistent with the assumption that the same set of highly constrained genes identified in ongoing genome-wide studies are associated with ASD, regardless of ancestry. This perspective also receives support from common variant studies in complex traits, where causal effects appear to be highly similar across ancestries (Hou *et al*., 2023). These observations are consistent with the neurobiology of ASD being universal and provide support for the translatability of clinical genetic approaches across ancestries. Using clinical genetics software platforms, we confirm the overall translatability of clinical genetic approaches when focusing on rare deleterious variation; however, we also reveal significant differences in the rate of P/LP variants between AMR and non-AMR individuals and EUR and non-EUR individuals.

The causes driving differences in rates of P/LP needs to be better understood. An recent study focusing on pediatric patients with significant neurologic, cardiac, or immunologic conditions reported similar diagnostic yield for genome sequencing in European Americans and Latin Americans (19.8% versus 17.2%); however, yields were lower (11.5%) and inconclusive results higher in African Americans (Abul-Husn *et al*., 2023). In that study, genome sequencing was carried out by commercial diagnostic laboratories, making use of a proprietary pipeline that incorporates variant databases; the degree to which proprietary algorithms and reliance on previously observed variation influenced the higher rate of inconclusive results cannot be determined. Our results suggest that with a focus on deleterious *de novo* variation, use of prior results are less necessary, and others have shown that even highly curated variant databases include false positive findings that can lead to incorrect information to subsequent families (Lek *et al*., 2016; Manrai *et al*., 2016; Sharo *et al*., 2023; Ciesielski *et al*., 2024).

Analysis of pathogenic variation in All of Us, which integrates data from a diverse cohort to identify genetic differences across ancestries, highlights the disparities in variant classification across populations. The study examined P/LP variants identified through WGS, showing differences as a function of ancestry, with 42% fewer pathogenic variants identified in Latin American versus European individuals (1.32% versus 2.26%) (Venner *et al*., 2024). All of Us analyses used Neptune, a system developed by the eMERGE Consortium for clinical genetic reporting (Eric *et al*., 2021). Neptune relies heavily on variants identified in prior curated data, which will bias the findings in diverse populations. Consistent with this, analyses of the GALA cohort using Neptune shows lower rates of findings compared to non-AMR samples.

Our results suggest paths to improve genetic testing results. Clearly, of key importance is to use allele frequency from all relevant populations, as we have done here. Next, where possible, we recommend minimizing reliance on previously reported pathogenic variants. And, finally, we should recognize the challenges inherent in ancestries beyond European and a few other commonly characterized populations. For instance, we have focused on *de novo* variants and their interpretation in AMR populations. Variants called *de novo* in our sample, and within subjects, are likely a mixture of true and false positives. For populations not deeply characterized for genetic variation, it is reasonable to expect elevation in the false positive rate, simply because we do not know the frequencies of variants therein and which variants are relatively more common. For this reason, more of the variation called *de novo* is likely to be inherited variation.

At the same time, it is possible that unknown genomic complexity, such as common structural variants (Collins *et al*., 2020; Jun *et al*., 2023; Liao *et al*., 2023) elevate false negatives within these populations – including genomic variation important for phenotypes like autism. The combination of these three quantities, true positives, false positives, and false negatives, determine the total variation we observe. Based on our results, which show similar patterns to those observed in EUR studies, we can conclude that the vast majority of our results arise from true positives. Nonetheless, we should not conclude that populations are all the same when it comes to calling *de novo* variation. Indeed, we can be confident they are not, given what we know about increased genetic diversity in African populations (Yu *et al*., 2002; Gomez, Hirbo and Tishkoff, 2014; Pereira *et al*., 2021; Yilmaz *et al*., 2021) and the impact that cryptic structural variation and singleton events have on the reliability of calling ultra-rare variation. Only through deeper genetic studies can we expect completely comparable results to those of EUR population samples.

## Methods

### Cohort Description

GALA comprises multiple sites from North, Central, and South America recruiting AMR participants for studies on the risk architecture of ASD. ASD diagnoses are based on expert clinical evaluations using DSM-5 criteria, incorporating all available data including standardized assessments. Participants can be any age. Individuals with a known genetic condition (*e.g.*, fragile X syndrome) are excluded from analyses. Once a diagnosis of ASD is confirmed, the individual and their parents contribute a sample (blood or saliva) for genetic analyses. If both parents are not available, collection of other biological family members is encouraged (siblings, grandparents, etc.). Participating sites generally also collect additional clinical and family history information. Details on GALA sites are found in **Supplemental Materials**.

### Ancestry Determination and Sample Level QC

Latin American samples analyzed in the current freeze include (i) GALA participants (some published in Fu *et al*.,(Fu *et al*., 2022)), (ii) non-overlapping AMR samples in the Autism Sequencing Consortium (ASC) and Simons Powering Autism Research (SPARK) (SPARK Consortium., 2018) reported in Fu *et al*. (Fu *et al*., 2022), and (iii) additional AMR samples from the new release of SPARK (iWESv2). The current freeze includes trio data from 14,359 AMR samples, including 4,450 affected individuals (609 from GALA, and 3,841 from ASC and the SPARK releases) and 1,459 typically-developing siblings, and case-control data from 267 cases and 801 controls.

For ancestry determination, each of three jointly called datasets, derived from unpublished GALA sequencing, Fu *et al*., and SPARK (iWESv2), was combined with the HGDP + 1KG subset of gnomAD (Karczewski *et al*., 2020) and Principal Component Analysis (PCA) was performed in the joint dataset after they had been restricted to 5,000 ancestry informative SNPs (Purcell *et al*., 2014). A Random Forest classifier was trained on the HGDP + 1KG subset using the first 10 Principal Components to infer ancestry for all samples. Non-AMR cases included any individuals with ASD in ASC and the two SPARK releases that did not meet our criteria for genetically-inferred AMR ancestry (28,818 parents, 13,030 probands and 4,749 typically-developing siblings).

Hail 0.2 was used to process the SPARK (iWESv2) and unpublished GALA joint-genotyped VCFs. Multi-allelic sites were split, variants were annotated using the Variant Effect Predictor (VEP) (McLaren *et al*., 2016) and low-complexity regions (https://github.com/lh3/varcmp/blob/master/scripts/LCR-hs38.bed.gz) were removed. Hail’s *pc_relate()* function was used to confirm reported pedigrees and identify duplicate samples within and between datasets, which were removed. Sex was imputed using the *impute_sex()* function and genotype filters were applied as described previously (Satterstrom *et al*., 2020) methodology to generate working datasets (**Supplemental Figure 1**).

### De novo Variants

Previously published *de novo* calls were extracted from the Supplementary Table 20 from Fu *et al. (Fu et al., 2022)* For the unpublished GALA and the SPARK (iWESv2) datasets, *de novo* variants were called using the *my_de_novo_v16()* function (https://discuss.hail.is/t/de-novo-calls-on-hemizygous-x-variants/2357/19) with variant frequencies from the non-neuro subset of gnomAD exomes v2.1.1 as priors. Potential *de novo* variants were dropped if they were present at a frequency greater than 0.1% within the non-neuro subset of gnomAD v2.1.1, gnomAD v3.1.2, in any subpopulation of these gnomAD datasets, or the dataset in which they were called. Variants were further excluded if they had *“ExcessHet”* in the Filters field, exhibited a proband allele balance < 0.3, or demonstrated a depth ratio < 0.3. Only “HIGH” or “MEDIUM” confidence variants were kept, with the medium-confidence calls limited to a maximum allele count in the dataset of 1. A single variant per person per gene was chosen, giving preference to variants with more damaging consequences. Samples were finally excluded if the count of coding *de novo* variants was significantly greater than expected.

|  | **GALA**<br>**(unpublished)** | **GALA**<br>**(published)** | **Fu *et al.* 2022**<br>**Non-GALA**<br>**AMR** | **SPARK**<br>**(iWESv2)**<br>**AMR** | **Total** |
| --- | --- | --- | --- | --- | --- |
| **Probands** | 440 | 545 | 1,459 | 2,006 | **4,450** |
| **Siblings** | 26 | 92 | 650 | 691 | **1,459** |
| **Parents** | 854 | 1,130 | 2,812 | 3,654 | **8,450** |
| **Cases** | 267 |  |  |  | **267** |
| **Controls** | 801 |  |  |  | **801** |
| **Total** | **2,388** | **1,767** | **4,921** | **6,351** | **15,427** |

### Inherited Variants

Starting with the same working datasets as for *de novo* calling, counts of transmitted and non-transmitted alleles were generated using Hail’s *transmission_disequilibrium_test()* function. Variants were filtered out if they were marked “*ExcessHet”* by GATK4 or had allele frequencies greater than 0.01% within their own dataset, within the non-neuro subset of gnomAD v2.1.1, gnomAD v3.1.2 or within any subpopulation of these gnomAD datasets. Variants with an allele count > 6 in the total parents of the dataset were excluded as well. Hard filtering was applied according to GATK recommendations (https://gatk.broadinstitute.org/hc/en-us/articles/360035890471-Hard-filtering-germline-short-variants). Final counts of transmitted and non-transmitted alleles were produced for PTV, MisB, MisA (1 ≤ MPC < 2) and synonymous variants.

### Case-control variants

Probands within incomplete trios were identified from the ASC and GALA cohorts and matched using the top 10 PCs (see Methods, Ancestry Determination) with non-psychiatric, unrelated controls from BioMe at a ratio of 3 controls to 1 case. Incomplete trios from SPARK (iWESv2) were removed. To ensure genome build standardization between these two cohorts, cram files from ASD cases were unmapped using GATK4 (Van der Auwera *et al*., 2013) and then remapped to a different version of the hg38 reference genome (https://biobank.ndph.ox.ac.uk/ukb/refer.cgi?id=838) using GATK3.5. Single Nucleotide Variants (SNVs) and insertions/deletions (indels) were joint-genotyped across cases using the Haplotypecaller of GATK4. Like for the trio dataset processing, Hail 0.2 (https://hail.is) was used to process the joint-genotyped VCF file. The *identity_by_descent()* function of Hail was used to test for relatedness, which resulted in the removal of thirteen cases. Sex was imputed for every sample using the *impute_sex()* function of Hail, and cross-checked with metadata provided by all sites to ensure sample concordance.

As was done for the previous datasets (Fu *et al*., 2022), multi-allelic sites were split, variants were annotated using the Variant Effect Predictor (VEP) and low-complexity regions were removed. Variants were removed if they had an allele count ≥ 2 in the entire case-control dataset, as well as an allele count ≥ 5 in the non-psychiatric subset of gnomAD v2.1.1. Genotype calls were filtered to genotype quality (GQ) > 25 and allele balance (AB) > 0.3. For case-control coverage harmonization, variants in high coverage, defined as a call rate ≥ 90%, were kept. To perform case-control matching, we excluded one case that was an outlier in the distribution of the number of synonymous variants. Finally, 267 cases were matched to 801 controls by sex and the first 10 Principal Components using the match_on function of the R package “optmatch” (https://github.com/markmfredrickson/optmatch).

### CNV Analysis

*De novo* CNVs called in Fu *et al*. (Fu *et al*., 2022) coming from AMR samples were extracted (1,861 probands and 680 unaffected siblings). Trio and case-control datasets were analyzed separately and GATK-gCNV (Babadi *et al*., 2023) was used to detect CNVs. First, raw cram files were compressed into read counts that covered the annotated exons to serve as input data. Then, a PCA-based approach that combines density and distance-based clustering was employed on the observed read counts to organize batches of samples for parallel processing. GATK-gCNV was run on cohort-mode analysis for 200 samples within the cluster identified through PCA analysis and the remaining samples were subjected to GATK-gCNV analysis using the case mode, with models specific to the cohort (368 probands and 29 typically-developing siblings). For quality control, CNV calls were processed according to Fu *et al. (Fu et al., 2022)* methodology; CNVs were retained if they had an allele frequency < 1% that spanned more than two captured exons. For homozygous deletions, the quality score (QS) threshold was set to the lesser of 400 or ten times the number of intervals. For heterozygous deletions, the QS threshold was set to the lesser of 100 or ten times the number of intervals. For duplications, the QS threshold was set to the lesser of 50 or four times the number of intervals. For sample-level quality control, samples were retained if the number of raw, autosomal CNV calls detected by GATK-gCNV did not exceed 200 and if the number of calls with QS ≥ 20 did not exceed 35. After quality control, 291 probands, 25 typically-developing siblings, 209 cases and 735 controls remained.

A gene was considered impacted by a deletion if at least 10% of its non-redundant exons were overlapped by the deletion. For a duplication, a gene was considered impacted if at least 75% of its non-redundant exons were overlapped. Additionally, CNVs were annotated against a list of 79 curated genomic disorder (GD) loci (see Fu et al. (Fu *et al*., 2022), Supplementary Table 10) and a CNV call was classified as a genomic disorder CNV if it shared at least 50% reciprocal overlap with an annotated GD.

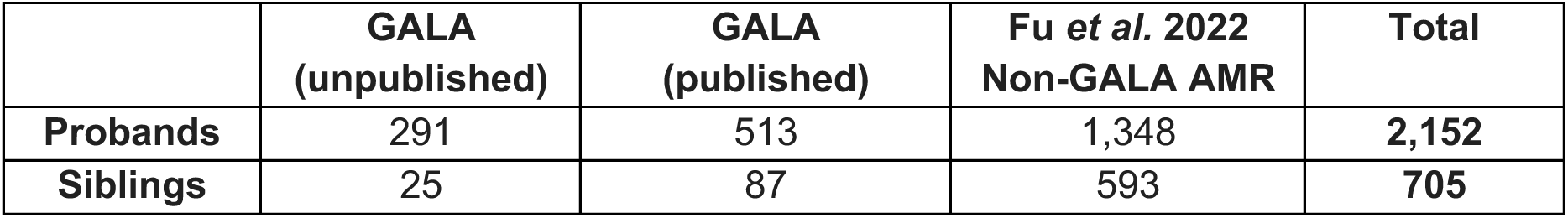
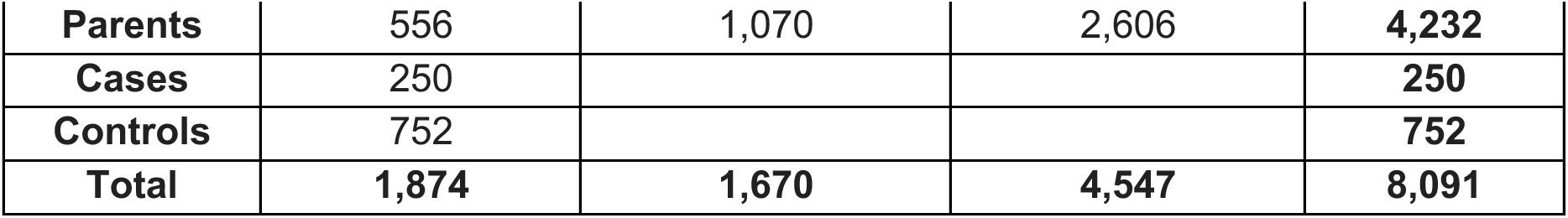

|  | **GALA**<br>**(unpublished)** | **GALA**<br>**(published)** | **Fu *et al.* 2022**<br>**Non-GALA AMR** | **Total** |
| --- | --- | --- | --- | --- |
| **Probands** | 291 | 513 | 1,348 | **2,152** |
| **Siblings** | 25 | 87 | 593 | **705** |
| **Parents** | 556 | 1,070 | 2,606 | **4,232** |
| **Cases** | 250 |  |  | **250** |
| **Controls** | 752 |  |  | **752** |
| **Total** | **1,874** | **1,670** | **4,547** | **8,091** |

### Genetic Association Analyses

TADA (Fu *et al*., 2022) was performed for 3 types of inheritance classes; *de novo* (PTV, MisB, MisA, DEL, DUP), inherited (PTV, MisB, MisA) and case-control (PTV, MisB, MisA, DEL, DUP) variation. CNVs resulting from Non-Allelic Homologous Recombination were excluded and only CNVs impacting fewer than nine constrained genes were retained (LOEUF < 0.6) (**Supplementary Tables 9-15**).

Bayes factors (BFs) were constructed separately for each variant class (PTV, MisA, MisB, DEL, DUP) as described, accounting for sample size and directly using relative risk priors from Fu *et al*. directly (see Fu et al. (Fu *et al*., 2022), Supplementary Table 8). Previously published mutation rates were adjusted to align with the observed mutation counts in unaffected siblings for each variant type in the dataset (Fu *et al*., 2022).

### ACMG Interpretation of Variants

*De novo* variants, including those in X-linked genes and autosomal genes excluded from the genetic association analyses due to the absence of mutation rates and/or LOEUF scores (**Supplementary Table 16**), were analyzed. In addition to applying the allele frequency cutoff of 0.1% described above (see Methods, *De novo* Variants), X-linked variants were subjected to an allele frequency cutoff of 0.1% in the male non-psychiatric subsets of gnomAD versions 2.1.1 and 3.1.2. and their subpopulations. This resulted in 20,571 *de novo* variants being included for analysis. In addition, inherited variants in a list of well-defined X-linked genes implicated in ASD and/or intellectual disability (**Supplementary Table 5**) were extracted from the datasets and subjected to the same allele frequency cutoff.

The commercially available VarSome package (Kopanos *et al*., 2018) was used to evaluate the clinical impact of both *de nov*o variants and X-linked inherited variation in the selected genes. Given the large number of variants, a batch environment was utilized, which limited the parameters that could be optimized for each gene. Additionally, as ACMG guidelines (Richards *et al*., 2015) consider patient phenotype, the focus was placed on genes for which there was a reported relationship with an ASD phenotype (Autism Spectrum Disorder, Autism, Autistic Behavior) and/or with a broader NDD phenotype (including the three ASD terms, as well as Intellectual Disability, Global Developmental Delay, Seizure, Epileptic Encephalopathy, and Complex Neurodevelopmental Disorder), without knowing the full spectrum of non-ASD phenotypes in the participants. Hence, the results presented here (**Supplementary Tables 6-7**), while based on a more transparent algorithm, should not be considered fully compliant with ACMG reporting guidelines.

The *api.batch_lookup* function in VarSome was used to obtain germline variant-level information related to ACMG classification, nucleotide substitution and amino acid substitution, along with pathogenicity predictions. When possible, transcripts with the most severe coding impact were selected. Otherwise, the MANE Select transcript, longest canonical transcript, MANE Plus transcript, longest transcript, or RefSeq transcript were chosen in that order by default.

For *de novo* variation, variant lists containing unique sets of variants found in each sex and zygosity were annotated. Inheritance in VarSome was set to “Confirmed De Novo.” Output from each list was returned in separate JSON files, which were then read into R for downstream processing into tab-separated tables. Inherited variation was examined in a similar manner, however, inheritance was set to the parent of origin of the variant.

To extend these analyses further, we used Neptune (Eric *et al*., 2021), examining 73 ACMG actionable genes analyzed in All Of Us (Venner *et al*., 2024). The VIP database used for annotation in Neptune was downloaded from https://gitlab.com/bcm-hgsc/neptune in VCF format and all variants were lifted over (Nassar *et al*., 2022) from GRCh37 to GRCh38. Clinical significance annotations were parsed from the INFO field and variants classified as Pathogenic/Likely Pathogenic, Unknown Significance and Benign/Likely Benign variants were noted. All rare variants in probands, regardless of mode of inheritance, were used in these analyses. Of the 73 genes, Veneer *et al*. annotated only biallelic variants as P/LP in 3 recessive genes (*MUTYH*, *ATP3B*, *KCNQ1)* and only a specific variant as P/LP in *HFE*; we did not observe P/LP variants in these 4 genes, so no additional corrections were made.

## Data Availability

All data produced in the present study are available upon reasonable request to the authors.

## Acknowledgments

GALA is currently supported by the National Institutes of Health grant MH128813 and by the Seaver Center. GALA originated with sites from, and with support of, the Autism Sequencing Consortium MH129724, MH111661, MH1000233. ASC analytical sites continue to support GALA studies MH129722 and MH115957. This work was supported in part through the computational and data resources and staff expertise provided by Scientific Computing and Data at the Icahn School of Medicine at Mount Sinai and supported by the Clinical and Translational Science Awards (CTSA) grant UL1TR004419 from the National Center for Advancing Translational Sciences. Research reported in this paper was also supported by the Office of Research Infrastructure of the National Institutes of Health under award numbers S10OD026880 and S10OD030463. The content is solely the responsibility of the authors and does not necessarily represent the official views of the National Institutes of Health.

This study makes use of data generated by the DECIPHER community. A full list of centres who contributed to the generation of the data is available from https://deciphergenomics.org/about/stats and via email from. DECIPHER is hosted by EMBL-EBI and funding for the DECIPHER project was provided by the Wellcome Trust [grant number WT223718/Z/21/Z].

## Author Information

### Author Contributions

K.R., B.D., C.B. and J.D.B. designed the study. T,L., T.P., C.R.S., C.M.C., J.L.A., G.S.C., H.C., R.C., C.I.S.C., M.L.C., A.D.P.L., M.F., E.F., L.G., A.C.D.E.S.G., A.J.G., L.C.H., N.L., Y.L., D.L.N., R.O., K.P.P., I.P., R.S., H.M.S., L.T., J.Y.T.W., L.A.-G., L.A.C., C.S.C.-F., I.H.-P., A.K., M.C.L., L.M., M.R.P.-B., M.A.P.-V, P.S., F.T., M.P.T., M.E.T., M.J.D. and J.D.B. contributed samples and generated data. J.D.B, B.D., B.M., S.D.R., L.K., L.S., J.M.F, F.K.S, S.J, and M.N.A developed methodology and performed analysis. J.D.B, B.D., C.B., E.H.C., T.L, L.S., and M.N.A wrote the manuscript.

## Supplemental Tables

An excel spreadsheet with 18 tabs is included with the following Tables

Supplementary Table 1 Pedigree file for all samples in the study

Supplementary Table 2 List of de novo variants used in TADA

Supplementary Table 3 List of CNVs used in TADA

Supplementary Table 4 TADA results

Supplementary Table 5 X-linked genes included in VarSome analyses

Supplementary Table 6 VarSome results matching phenotypes in Autism Spectrum Disorder, Autism, or Autistic Behavior

Supplementary Table 7 VarSome results matching phenotypes in Autism Spectrum Disorder, Autism, Autistic Behavior, Intellectual Disability, Global Developmental Delay, Seizure, Epileptic Encephalopathy, or Complex Neurodevelopmental Disorder

Supplementary Table 8 Yield of results for VarSome and for Neptune

Supplementary Table 9 TADA counts for new GALA samples

Supplementary Table 10 TADA counts for AMR samples in ASC as reported in Fu (including published GALA samples)

Supplementary Table 11 TADA counts for AMR samples in SPARK as reported in Fu et al

Supplementary Table 12 TADA counts for AMR samples from SPARK-iWESv2

Supplementary Table 13 TADA counts for FuCOMP (ASC) samples

Supplementary Table 14 TADA counts for FuCOMP (SPARK) samples

Supplementary Table 15 TADA counts for new GALA case-control samples

Supplementary Table 16 List of genes included in VarSome (with a flag to indicate inclusion in TADA analyses)

Supplementary Table 17 Enrichment analysis of 35 GALA genes for GO Biological Processes

Supplementary Table 18 Enrichment analysis of 35 GALA genes for Mammalian Phenotypes.

**Supplemental Figure 1.**
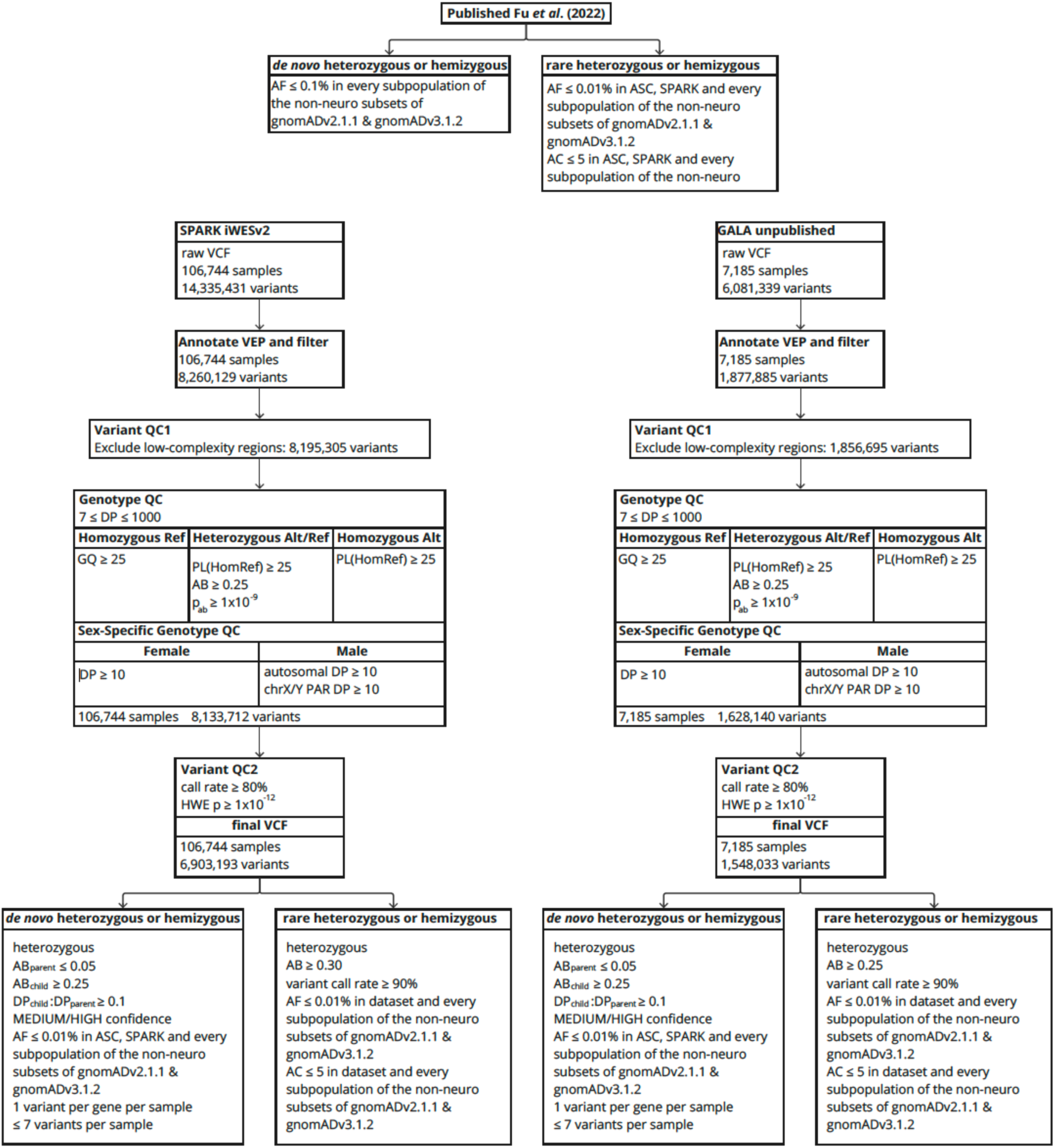
Data processing for samples from three different data sources. The figure describes the variant, genotype, and sample quality control steps that were implemented to process the raw, joint-genotyped VCFs and generate the de novo and inherited calls used for downstream analyses. Sample counts are tabulated before downstream ancestry filtering.

**Supplemental Figure 2.**
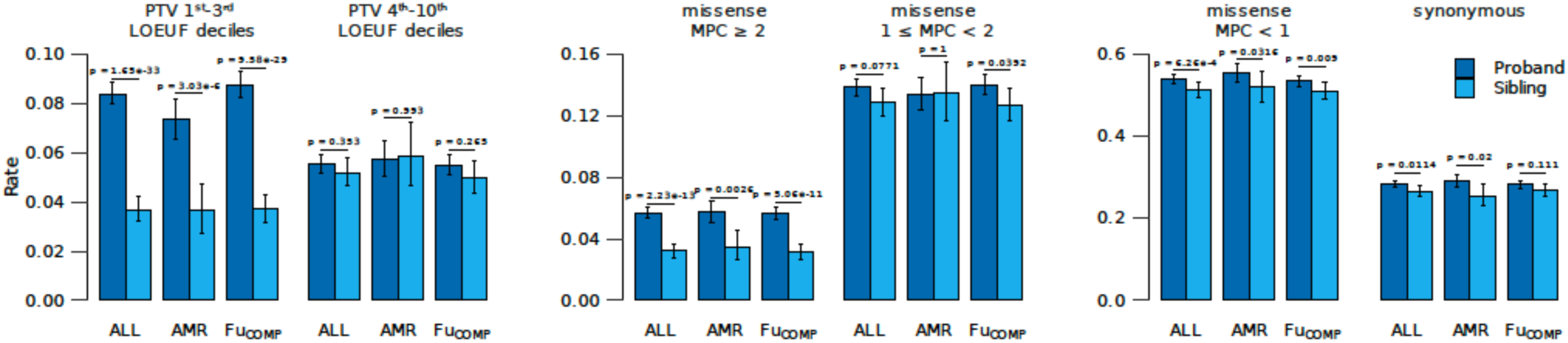
Comparison of rare de novo variant counts per sample between ASD probands and unaffected siblings across different ancestries in the current analysis. The average number of rare variants per sample is compared between ASD probands (dark blue) and their unaffected siblings (light blue) for all ancestries (ALL: 17,480 probands and 6,208 siblings), Admixed American (AMR: 4,450 probands and 1,459 siblings), and non-Admixed American (Fu_COMP_: 13,030 probands and 4,749 siblings). The analysis considers: (**left**) protein truncating variants (PTVs) in highly constrained genes (LOEUF deciles 1–3, 5,363 genes) and less constrained genes (LOEUF deciles 4–10, 12,765 genes); (**middle**) missense variants categorized by predicted functional severity (MPC ≥ 2 for high severity, 1 ≤ MPC < 2 for moderate severity), and (**right**) MPC < 1 (for low severity) and synonymous missense variants. Error bars represent 95% confidence intervals.

**Supplemental Figure 3.**
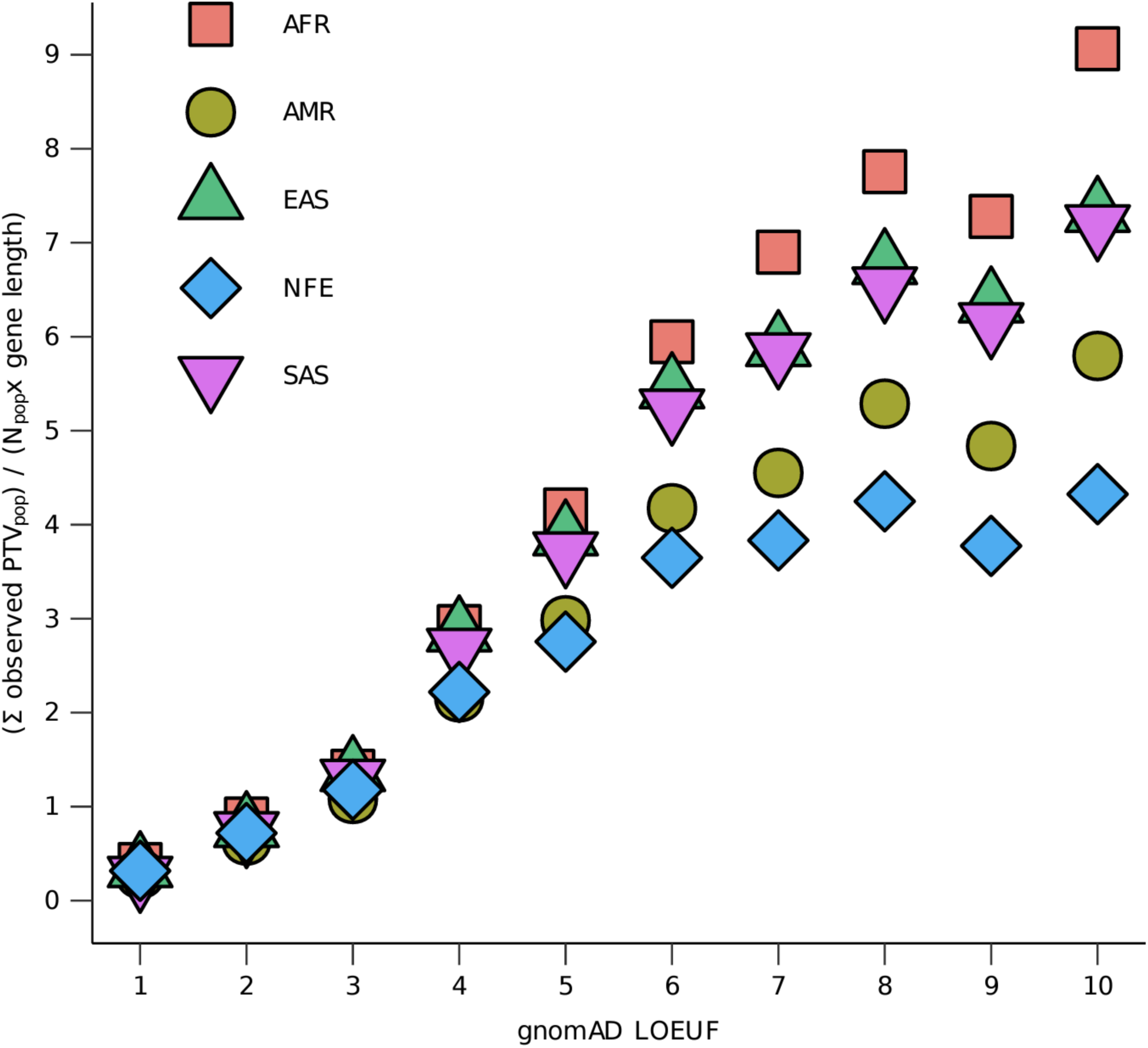
Ratio of observed PTVs across different ancestries in gnomAD. The sum of observed protein truncating variants (PTVs) per ancestry is plotted, scaled to each population’s size and total gene coding sequence length within gnomAD LOEUF deciles. The plot includes African/African American (AFR, red, N=8,128), Admixed American (AMR, green, N=17,296), East Asian (EAS, purple, N=9,197), Non-Finnish European (NFE, blue, N=56,885), and South Asian (SAS, yellow, N=15,308) ancestries. The population sizes in gnomAD v2.1.1 vary across ancestries, with larger sample sizes for some groups. LOEUF deciles represent levels of gene constraint, with lower deciles indicating more constrained genes.

**Supplemental Figure 4.**
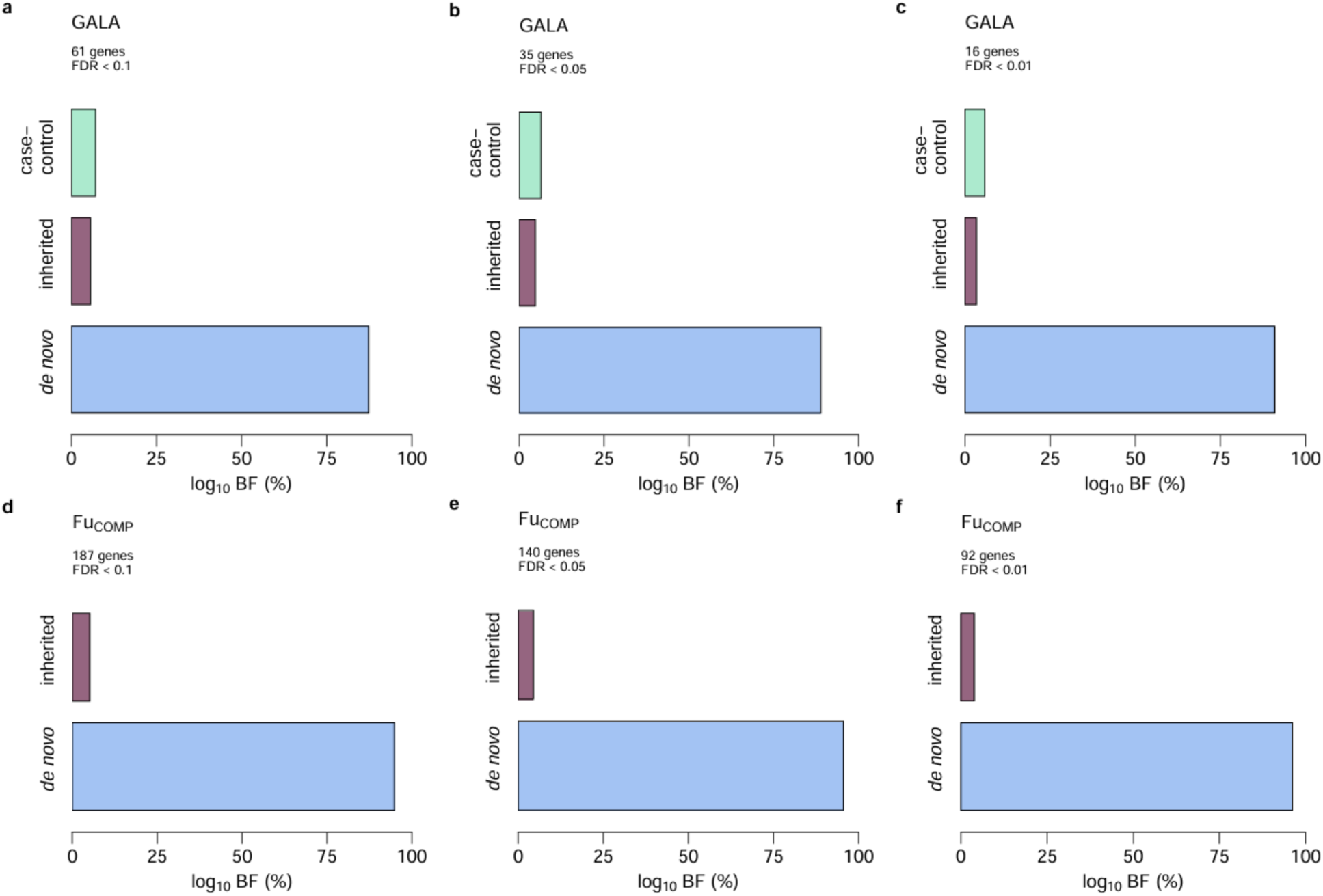
Relative contribution to TADA signal by mode of inheritance. The proportional impact of each inheritance mode on the ASD-associated genes is shown at three false discovery rate (FDR) thresholds: ≤ 0.1 (**a, d**), ≤ 0.05 (**b, e**), and ≤ 0.01 (**c, f**). Panels (**a–c**) display results for the GALA cohort, while panels (**d–f**) show results for the Fu_COMP_ subset from Fu *et al*. (2022).

**Supplemental Figure 5.**
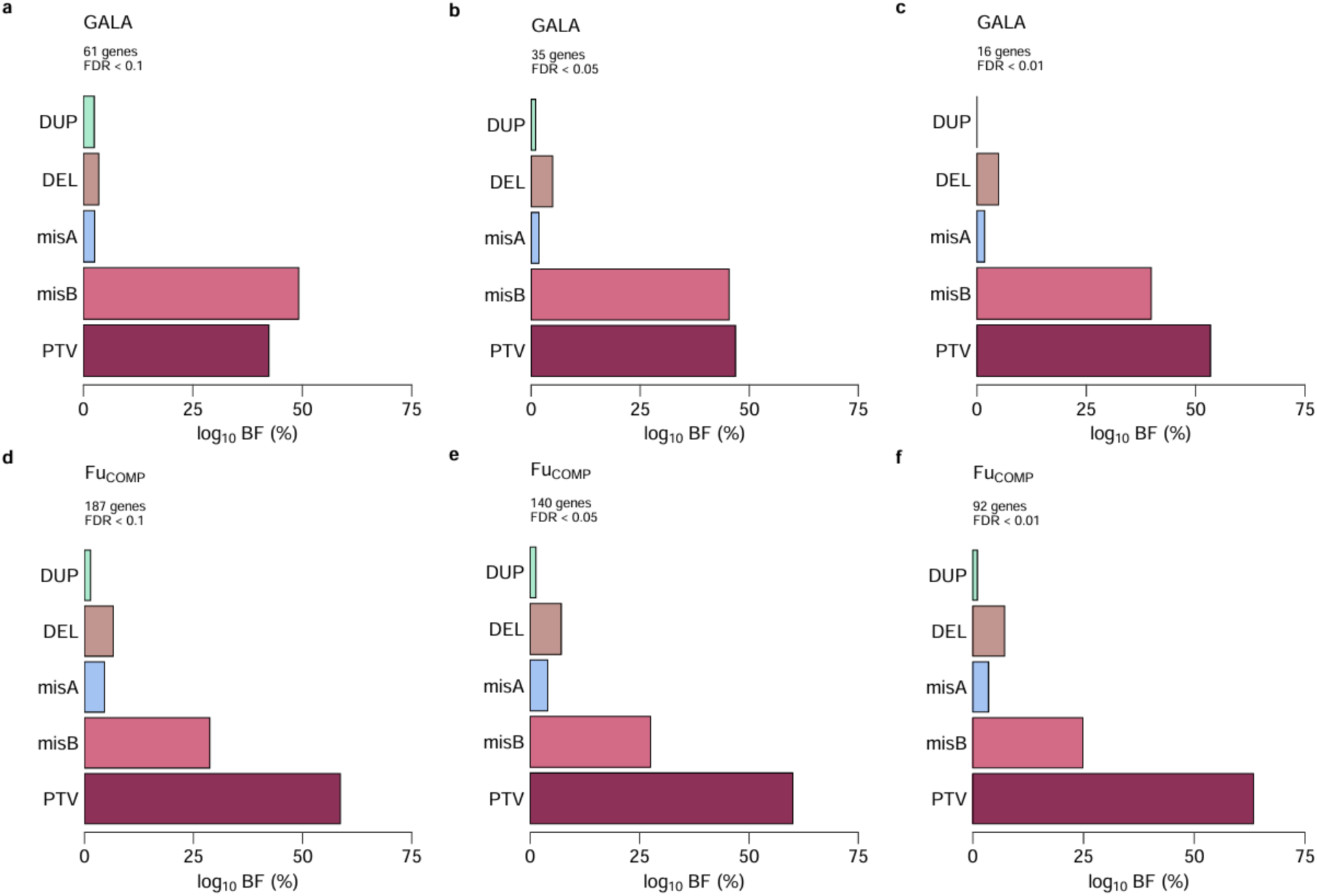
Relative contribution to TADA signal by variant type. The proportional impact of each variant type on the ASD-associated genes is shown at three false discovery rate (FDR) thresholds: ≤ 0.1 (**a, d, g**), ≤ 0.05 (**b, e, h**), and ≤ 0.01 (**c, f, i**). Panels (**a–c**) display results for the GALA cohort, while panels (**d–f**) show results for the Fu_COMP_ subset from Fu *et al*. (2022).

**Supplemental Figure 6.**
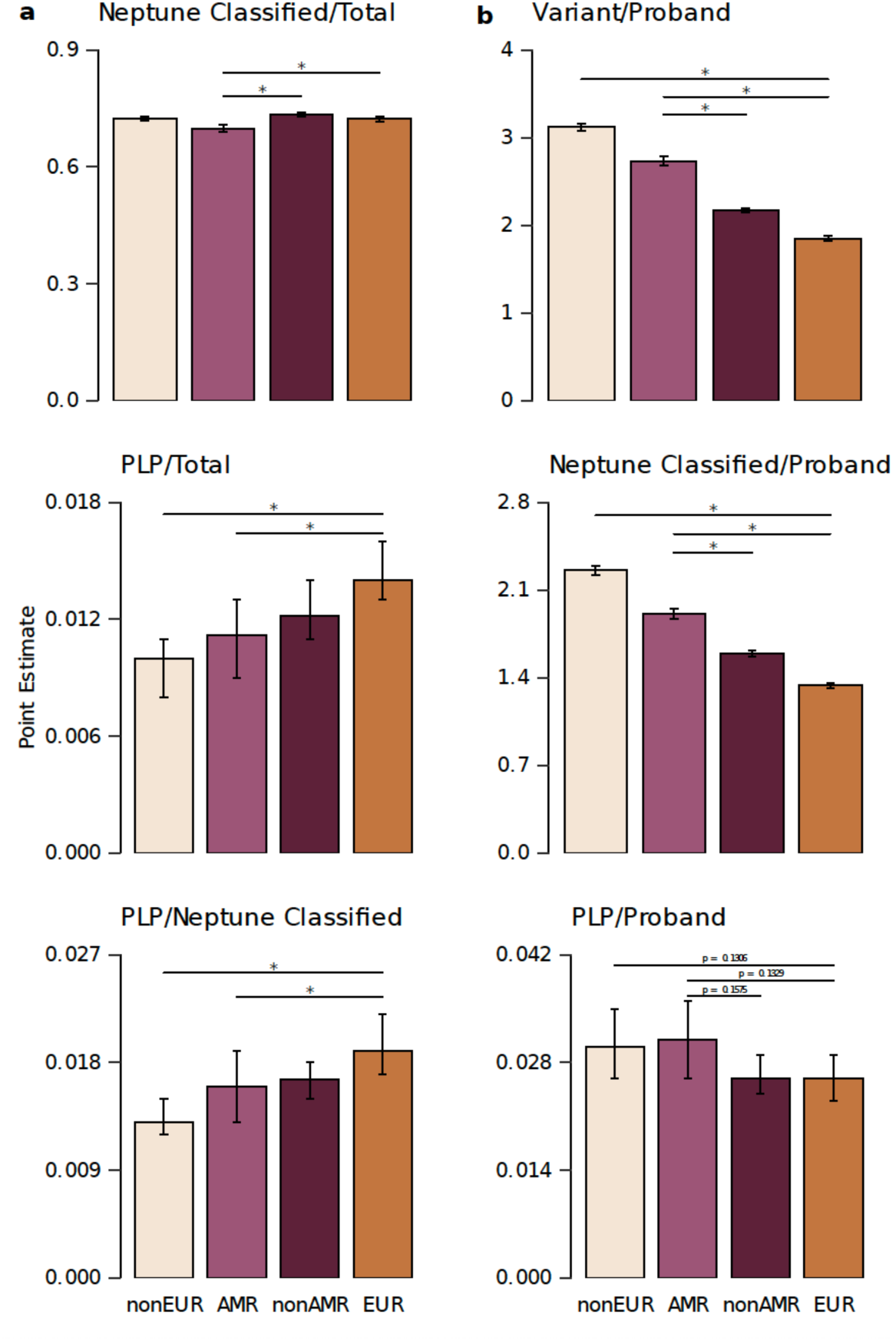
Classification rates and proportions of P/LP variants across AMR and non-AMR populations using Neptune. The figure compares the classification rates and proportions of pathogenic/likely pathogenic (P/LP) variants in the indicated subsamples. (**a**) The ratio of (**upper**) classified variants (by Neptune) to total variants, (**middle**) P/LP variants to total variants, and (**lower**) P/LP variants to Neptune classified variants is shown for AMR, non-AMR, non-European (non-EUR) and EUR ancestries. (**b**) Comparisons include (**upper**) the total number of variants, (**middle**) the number of classified variants, and (**lower**) the number of P/LP variants, all expressed per proband. AMR participants have more variants per individual (both total and Neptune-classified) compared to non-AMR participants, but a reduced ability of Neptune to classify variants in AMR contributes to a slightly lower proportion of P/LP variants per individual. Similar results are seen for non-EUR versus EUR. Error bars represent 95% confidence intervals. Statistical significance of differences is indicated by p-values.

**Supplemental Figure 7.**
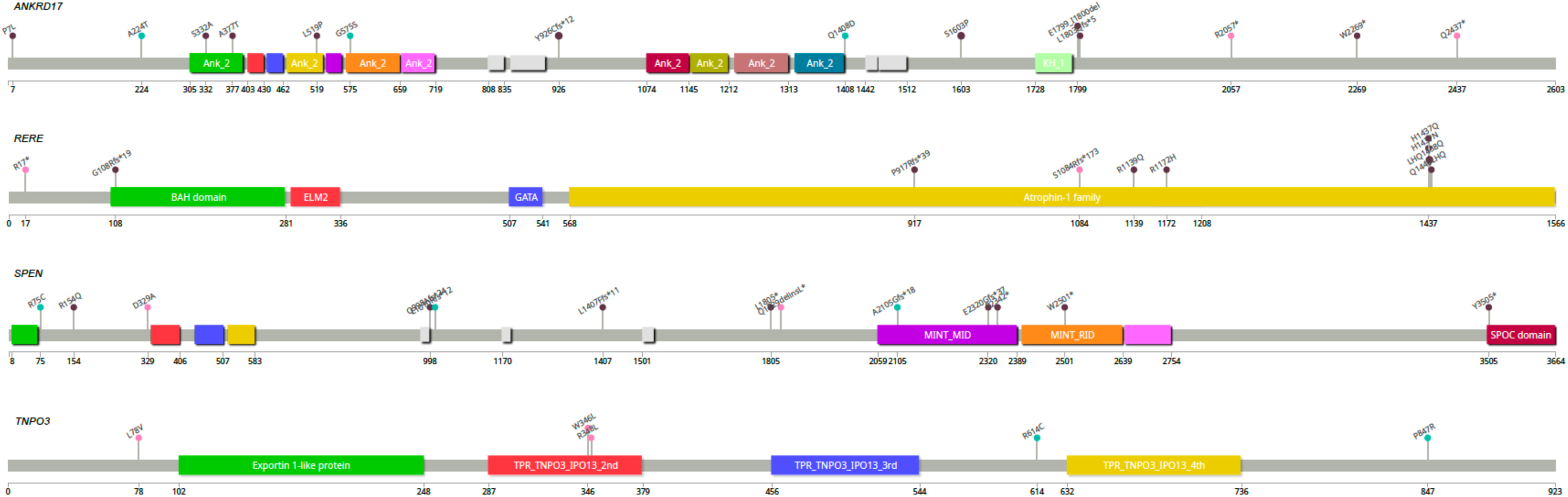
“Lollipop” diagrams illustrating variants identified in ASD-associated genes. Variants observed in GALA analyses of AMR individuals are marked with pink circles, those found in Fu_COMP_ individuals are marked with green, and variants found in DECIPHER are in purple. Figures were generated using the ‘lollipop’ software package (Jay and Brouwer, 2016)

